# How do we explain painful chronic non-traumatic knee conditions to children and adolescents? A multiple-method study to develop credible explanations

**DOI:** 10.1101/2022.12.15.22283510

**Authors:** C Djurtoft, MK Bruun, H Riel, MS Hoegh, B Darlow, MS Rathleff

## Abstract

**INTRODUCTION:** Perceived diagnostic uncertainty can leave adolescents confused about their condition and impede their ability to understand *“what’s wrong with me”*. Our aim is to develop credible explanations (trustworthy and understandable explanation of the condition) for adolescents suffering from chronic non-traumatic knee pain.

**METHODS:** This multiple-method study integrated findings from a systematic literature search of qualitative studies, an Argumentative Delphi with international experts (n=16) and think-aloud sessions with adolescents (n=7). Experts provided feedback with arguments on how to communicate credible explanations to meet adolescents’ needs. We analyzed feedback using thematic analysis before tailoring explanations to end-users.

**RESULTS:** We screened 3.239 titles/abstracts and included 16 papers which explored diagnostic uncertainty from adolescents’ and parents’ perspectives. Five themes emerged: (1) Understanding causes and contributors to the pain experience, (2) Feeling stigmatized for having an invisible condition, (3) Having a name for pain, Controllability of pain, and (5) Worried about something being missed. The Argumentative Delphi revealed the following themes: (1) Multidimensional perspective, (2) Tailored to adolescents, (3) Validation and reassurance, and (4) Careful wording. Merging findings from the systematic search and the Delphi revealed three essential domains to address in credible explanations: “What is non-traumatic knee pain and what does it mean?”, “What is causing my knee pain?” and “How do I manage my knee pain?”.

**CONCLUSIONS:** Six credible explanations for the six most common diagnoses of chronic non-traumatic knee pain were developed. We identified three domains to consider when tailoring credible explanations to adolescents experiencing non-traumatic knee pain.

## 1. Introduction

Chronic musculoskeletal pain (CMP) is recognized as a potential threat to the health of adolescents due to negative effects on physical, psychological, and social domains of life [25,26,32,55]. CMP can affect multiple body parts and areas, with knee pain being the most common site of pain in adolescents [19,55]. In recent years, research has uncovered the importance of how we communicate with people who have pain and the risks associated with many labels that are used for pain complaints [14,18]. However, communicating credible explanations beyond diagnostic labels remains a significant challenge for clinicians [14,46,48]. These issues have resulted in a wave of research agendas, where especially diagnostic uncertainty (*defined as “…subjective perception of an inability to provide an accurate explanation of the patient’s health problem”*) has gained a lot of attention lately [3,9,11,52].

Perceived diagnostic uncertainty can leave adolescents confused and impedes their ability to understand *“what’s wrong with me”* [23,46,62]. Uncertainty may have a cascading effect of higher anxiety, pain catastrophizing, fear of pain and lower acceptance of self-management recommendations [30,46,62]. The diagnostic process has a high impact on how the diagnosis of CMP is communicated and understood by clinicians, adolescents, and parents [11,43,46,48]. Adolescents and parents often report that they do not understand explanations given by clinicians, why some tests are (or are not) conducted, or the diagnostic label itself [23,43,46]. Diagnostic uncertainty may result in an extended journey through the healthcare system during which both adolescents and their parents are troubled by the feeling of ‘something missing’ [10,46]. Consequently, many adolescents experience multiple referrals, searching for the ‘right’ diagnosis and a credible explanation for their pain [10,40,43].

Chronic non-traumatic knee pain negatively impacts adolescents’ quality of life, sleep quality, sports participation, and social interactions [19,25,53]. These non-specific conditions are often viewed as self-limiting by clinicians and pain-dismissing statements, such as “*just growing pains”* or *“it’ll go away”*, have been reported [54,62]; Adolescents living with chronic pain may also experience pain-related stigma from family members, and peers [17,64]. In contrast, recent research shows that about 50% of adolescents with knee pain will continue to have pain after two years [53]. Unlike traumatic knee pain, the clinical assessment in non-traumatic knee pain often relies on subjective tests, such as pain provocation or movement-based tests [7]. This lack of diagnostic clarity often causes perceived diagnostic uncertainty during the clinical encounter [11,41,46].

Lancet Child & Adolescent Health Commission have emphasized fundamental gaps in current management and research for pediatric chronic pain and highlighted that the main goal is to ‘make pain understood’ [14]. How patient education can be improved to help individuals become more knowledgeable of their condition was recently ranked as a top research priority in CMP [14,36,51]. To date, no studies have integrated findings from multiple perspectives (i.e., best research evidence, clinical expertise, and lived experience with adolescent knee pain) to develop credible explanations for chronic pain conditions. Therefore, this study aims to develop credible explanations for adolescents suffering from chronic non-traumatic knee pain to encompass for diagnostic uncertainty.

## 2. Method

This multiple-method study was conducted through an iterative process, utilizing qualitative and quantitative methods (Figure 1). There were three steps: 1) two systematic literature searches, both aiming to answer, *“what information needs to be included in a credible explanation?”*, 2) An argumentative Delphi process to explore *“what do expert clinicians consider important in a credible explanation?”* and 3) think-aloud exercises (user testing) with adolescents to explore end-user perspective and ensure that the explanations being developed were in alignment of their needs and appear credible.

**Fig. 1.**
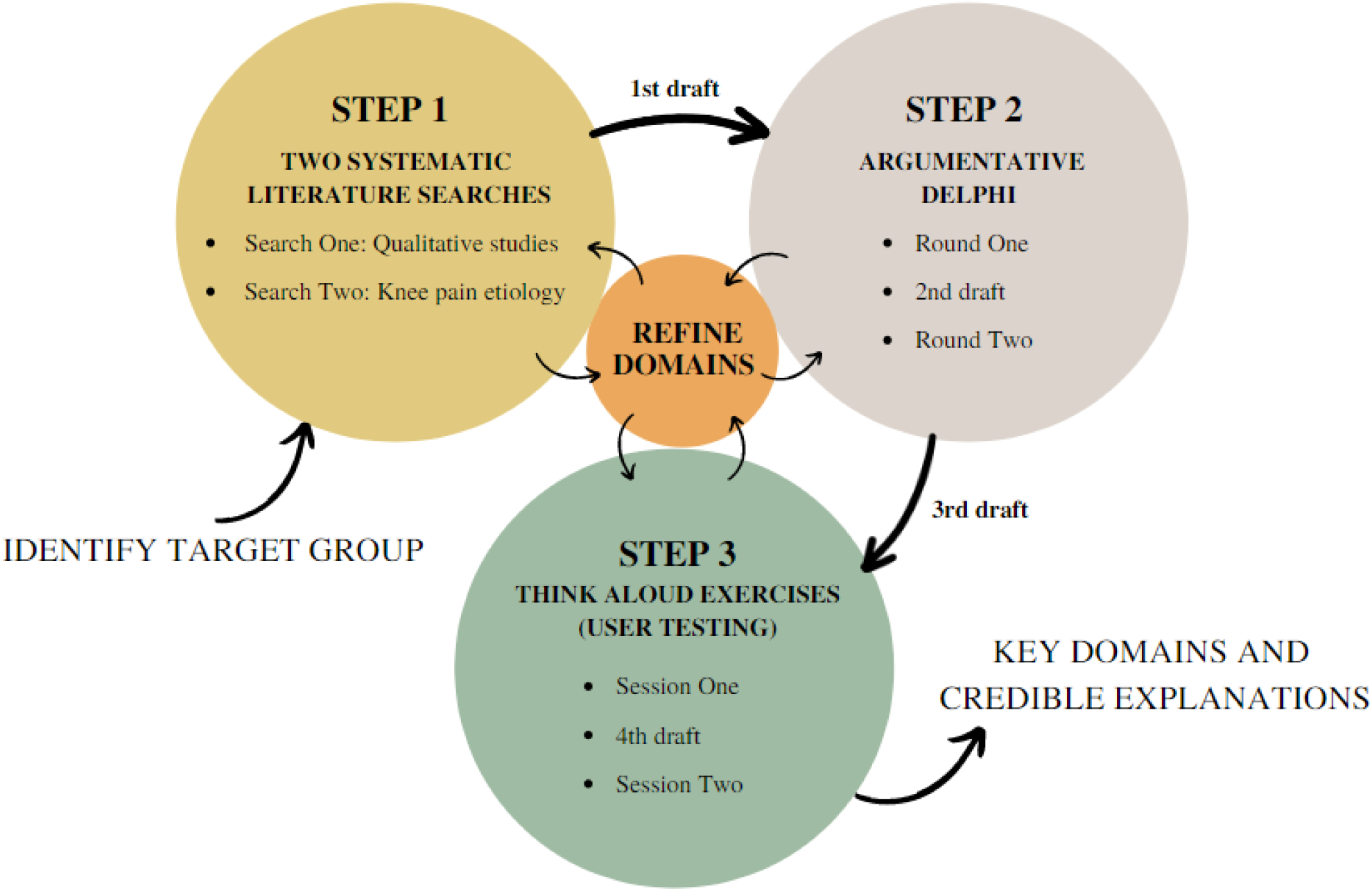
Multiple-method study design: Iterative process.

### 2.1. Step 1 – Two systematic literature searches

The evidence synthesis was based on data from two independent systematic literature searches in MEDLINE (via PubMed) in March 2022. The initial search (Search One) included papers of adolescents’ and parents’ information needs for understanding chronic primary musculoskeletal pain [49]; subsequently, a search (Search Two) was conducted to explore the etiology of non-traumatic knee pain among adolescents. Full description of both systematic searches and search strategies is available in *Appendix 1*.

#### 2.1.1. Development of credible explanations – From evidence synthesis to explanation

Using data collected from the two systematic searches (themes identified in the qualitative synthesis and synthesis of knee pain etiology) we created the first draft of a credible explanation for six of the most common non-traumatic knee pain conditions (Growth pain, Iliotibial Band Syndrome, Osgood Schlatter, Patellar Tendinopathy, Patellofemoral Pain, and Sinding-Larsson-Johansson) [21]. Existing wordings from leaflets and articles aimed at both the target population and adults were used as inspiration to build the first draft (Broad search on Google and Google scholar as well as leaflets known to the author group). The explanations were written in second person using lay language in a conversional style e.g., “your pain is most likely…” to enhance relatability for the adolescents. Any disagreements were resolved by discussion among the authors. The explanations were similar in terms of content with some minor diagnosis-specific alterations; for pragmatic reasons, we decided to only use the explanation of Osgood-Schlatter disease and applied all changes to the rest of the diagnoses afterwards.

### 2.2. Step 2 – Argumentative Delphi

We conducted a two-round Delphi survey procedure to gain input from experts in CMP and further explore and develop key domains of importance to include in a credible explanation (*Appendix 2*). The Delphi survey was piloted on two participants outside the author group to ensure readability, usability, and functionality [13].

#### 2.2.1. Recruitment of experts in musculoskeletal pain

We defined experts in musculoskeletal pain as being healthcare professionals (+5 years of clinical experience) currently working with adolescents (defined by the World Health Organization as the second decade of life, 10-19 years old [59]) experiencing musculoskeletal pain and/or working in the field of CMP research. International experts were recruited through multimodal recruitment strategy: personal networks (e.g., email) and advertisement on social media *(Facebook, LinkedIn, and Twitter)*. The social media post had a survey-link to REDCap (Research Electronic Data Capture), used as initial registration for experts interested in participating in the survey *“development of credible explanations” (Appendix 3)*.

Experts were first asked to complete demographic data (nationality, profession, years of clinical experience, area of expertise, and current occupation) and were then asked to select their diagnosis of expertise (multiple options were allowed). We aimed for a minimum of 15 experts from different healthcare professions (e.g., psychologists, physiotherapists, medical doctors, etc.) to ensure comprehensive and diverse opinions, reduce selection bias and allow for potential dropouts [50,63].

#### 2.3.2. Survey procedure and data analyses: Argumentative Delphi

Experts were sub-grouped into their self-reported area of expertise and received a link to the credible explanation for these conditions (e.g., Osgood-Schlatter disease and Patellofemoral Pain) via a REDCap survey. Experts only received credible explanations for diagnoses within their field of expertise and a maximum of two diagnoses each. First, experts were asked to read and rate the overall impression of the credible explanation. We used a five-point Likert scale *(Strongly disagree to Strongly agree)* for three questions to measure the degree of perceived diagnostic uncertainty adapted from Pincus et al *(Appendix 4)* [52]. Secondly, experts were asked to suggest changes to the credible explanations in a downloadable Microsoft Word-document and provide arguments and reasons for their suggestions. Any changes without reasons were not considered.

After the completion of Round One, all arguments and reasons (e.g., “*Important to emphasize how validation is key*”) were collated and analyzed using reflexive thematic analysis in accordance with Braun and Clarke’s six phases [4,5]. CD and MKB read and reread all feedback and arguments to familiarize themselves with the data before independently coding arguments and reasons to identify themes. Potential themes were noted in a coding list and subjected to iterative review. Refined themes were generated iteratively in accordance with the phases in reflexive thematic analysis. We chose not to rank arguments (as is common with Delphi processes) or use computerized methods, but to evaluate arguments equally using qualitative thematic analysis, allowing for a deeper understanding of our data [20,58,63].We merged themes from qualitative literature search and Round One of the Delphi to ensure all essential perspectives were included in our model of the final domains.

These domains were used as subheadings to re-structure the revised drafts of the credible explanations (second draft) for Round Two of the Delphi. The whole data analysis and revised version underwent review during a meeting in the author group that aimed to reach consensus on changes, revisions, and structure of the revised (second) draft. After the group meeting, the revised six explanations were sent out to all authors for final approval.

Round Two used the same procedure as described in Round One and aimed to collect feedback and ratings on the second draft of the credible explanations and provide stability of results (i.e., Likert-rating did not decrease) [20]. Experts were asked to rate the overall impression of the second draft of the credible explanation, using the same Likert scale as the first round and further provide additional feedback in the survey comment section; we aimed for a response rate of ≥80% [63]. A two-week deadline was chosen to ensure participants had enough time to attend and at the same time short enough to maintain their interest and minimize attrition [50,63]. Reminders were sent out twice (after week one and week two) if no response was received.

### 2.3. Step 3 – Think-aloud exercises (User testing)

In the last step, we used an iterative design consisting of user testing, feedback, input from the author group, and regular revisions, as recommended for the think-aloud method [8,27]. First, the credible explanations were translated into Danish to fit participants’ first language. Two bilingual *(English/Danish)* research assistants outside the author group made translations on all explanations independently of each other. Subsequently, we incorporated both translations to finish the third draft of the credible explanations before starting think-aloud exercises.

#### 2.3.1. End user involvement

Participants were recruited by purposive sampling using the authors’ social network. The think-aloud method was chosen because it utilizes simulation to learn about end-users’ perceptions, interactions, reasoning, and latent difficulties connected to artifact use, and then applies this knowledge to improve the design [8,35]. The interview guide was pilot tested internally in the author group prior to the first interview [31]. We planned two separate sessions including adolescents (at least three in each session to ensure a variety of thoughts as recommended) with and without non-traumatic knee pain and ages ranging from 8-17 years [8,27]. With specific emphasis to Faulkner’s observations on how 5-8 users will detect 85-95% of all usability problems based on the law of diminishing returns, participants were intentionally recruited according on perceived information power [16,37]. We chose to include the perspectives from one participant younger than the target group (<10 years old) based on the assumption that input from lower reading levels would enhance comprehension for older participants as well. Participants completed a demographic questionnaire: sex, age, current knee pain (yes/no) and knee pain duration. Think-aloud exercises were conducted face to face and performed by MKB (Audio recording with Dictaphone, *Olympus VN-711PC*) or online via Microsoft Teams (video recording) and transcribed non-verbatim for meaning retention as described by Kvale and Brinkmann [33].We chose non-verbatim for three reasons. First, our aim was to identify comprehension issues only; second, we did not intent to conduct an in-depth thematic analysis and third, we considered the exclusion of all unnecessary speech would make the transcript more informative for us to implement the suggestions provided by our participants [22]. Participants were instructed to verbalize thoughts as they occurred and say whatever came to mind as they read the credible explanation. The researcher reminded the participants to keep talking if they fell silent. The aim was to identify any discrepancies between the participants’ perception of the text compared and the intended meaning. Field notes were collected and then updated by reviewing the video recordings after the interview to capture missing points.

#### 2.3.2. Data analysis – Iterative revisions

All changes were based on notes and non-verbatim transcriptions. After Session One, revisions were made on a group meeting in the author group based on participants’ input during interviews (e.g., confusing phrases, or suggestions for improvement), before starting Session Two. The same procedure was used for the second session. We continued this iterative process until input for revisions were at a minimum, as recommendations prescribe [8,27].

### 2.4. Statistical analysis

All data from Argumentative Delphi were exported from the REDCap survey and through Microsoft Excel. Descriptive summaries for the Argumentative Delphi were reported for participant demographic characteristics, response rates for each survey round, and withdrawals. Data from think-aloud exercises underwent the same procedure. Results from five-point Likert scale were ranked in Microsoft Excel and visually interpretated; this allowed us to measure the experts perceived diagnostic uncertainty of the credible explanations between revisions.

## 3. Results

### 3.1. Two systematic literature searches

#### 3.1.1. Search One: Systematic search of qualitative studies

The systematic search of qualitative studies (Search One) yielded 3.239 papers. After screening the titles and abstracts, 26 papers underwent full-text review; we included 16 papers (*Appendix 5*). We organized concepts through abstraction into five themes: (1) Understanding causes and contributors to pain experience [12,17,24,29,30,32,39,43,45,46,60,64], (2) Feeling stigmatized for having an invisible condition [12,17,29,32,39,42,46,60,64], (3) Having a name for pain [6,29,42,43,46,60,61,64], (4) Controllability of pain [24,30,32,45,61,64], and (5) Worried about something being missed [6,39,43,46] (*Appendix 6)*. Six studies included parents, with no new themes identified by this group alone [6,29,42,43,46,64].

#### 3.1.2. Search Two: Systematic search of etiology for non-traumatic knee pain

The systematic search of etiology for non-traumatic knee pain (Search Two) yielded 2.934 papers. After screening the titles and abstracts, 112 underwent full-text review; we included 64 papers that described the etiology and/or pathogenesis of non-traumatic knee pain (*Appendix 5*).

### 3.2. Argumentative Delphi - Expert review

The Argumentative Delphi was conducted over two rounds between April 2022 and June 2022. Thirty-two participants signed up for taking part in the survey. Of these, 18 responded in the first round and 16 responded in the second round *(89% response rate)*. Two participants did not respond after inclusion and provided no reason for withdrawal. Therefore, 16 participants took part in this Argumentative Delphi survey (Table 1). Participants’ clinical experience ranged from 5 to 15+ years, with most >10 years. Seven countries were represented: Denmark (n=5), Turkey (n=4), United Kingdom (n=3), Canada (n=1), Ireland (n=1), Netherlands (n=1), Spain (n=1).

**Table 1.**
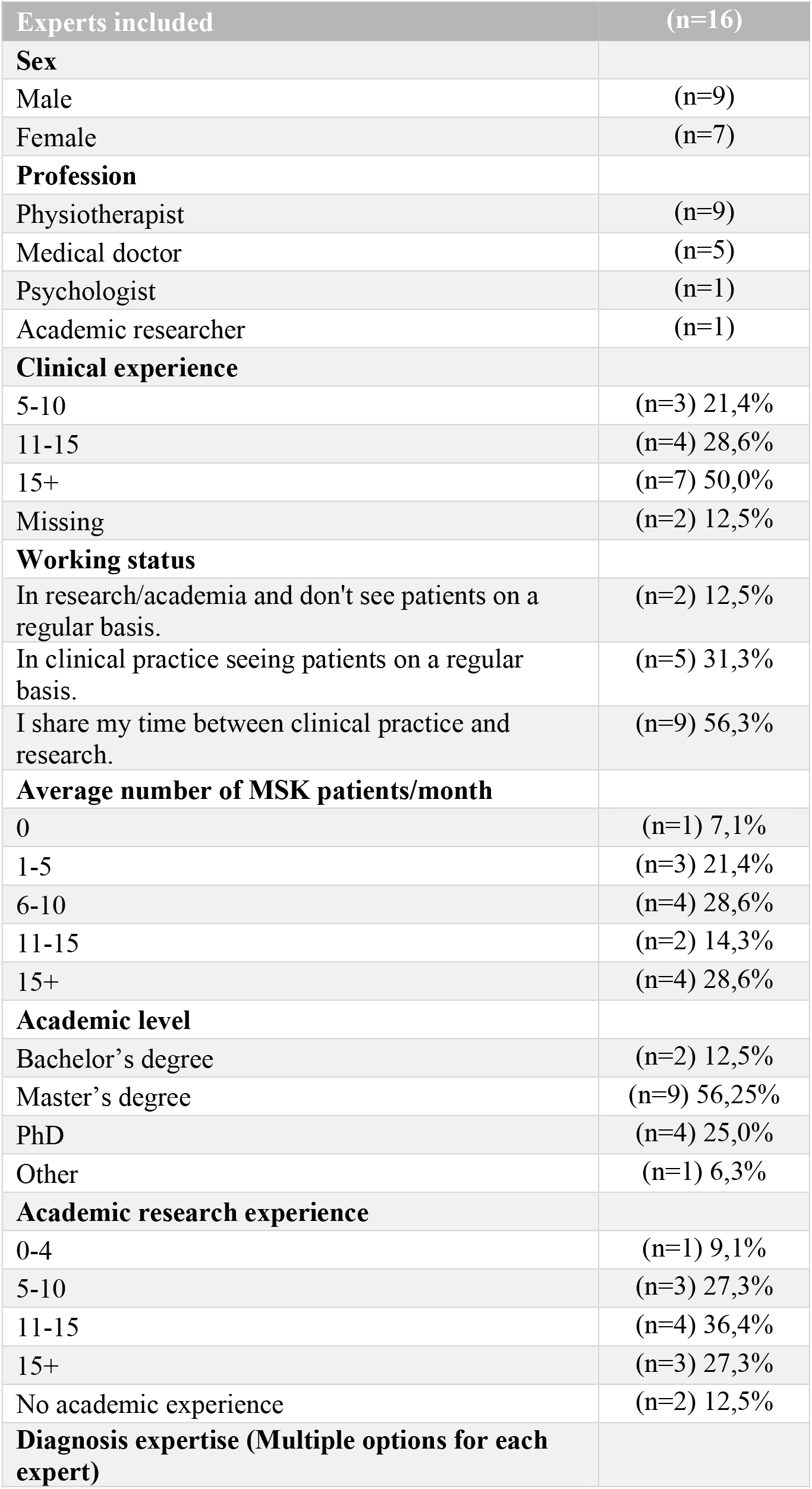
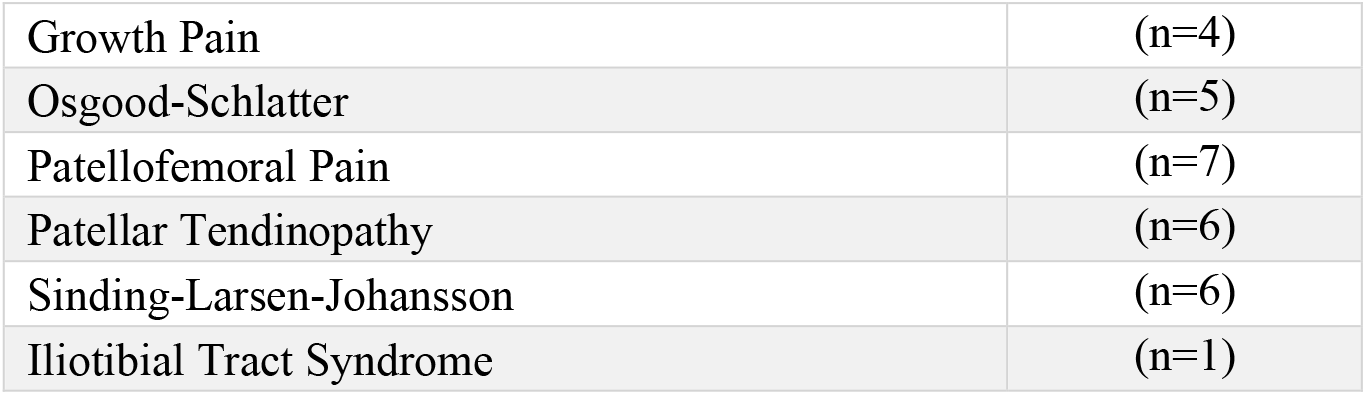
Characteristics of experts: Argumentative Delphi.

#### 3.2.1. Survey: Argumentative Delphi

Responses from Delphi Round One were collated and analyzed, from which 4 themes and 14 sub-themes emerged. Themes included: (1) Multidimensional perspective, (2) Tailored to adolescents, (3) Validation and reassurance, and (4) Careful wording (Figure 2).

**Fig. 2.**
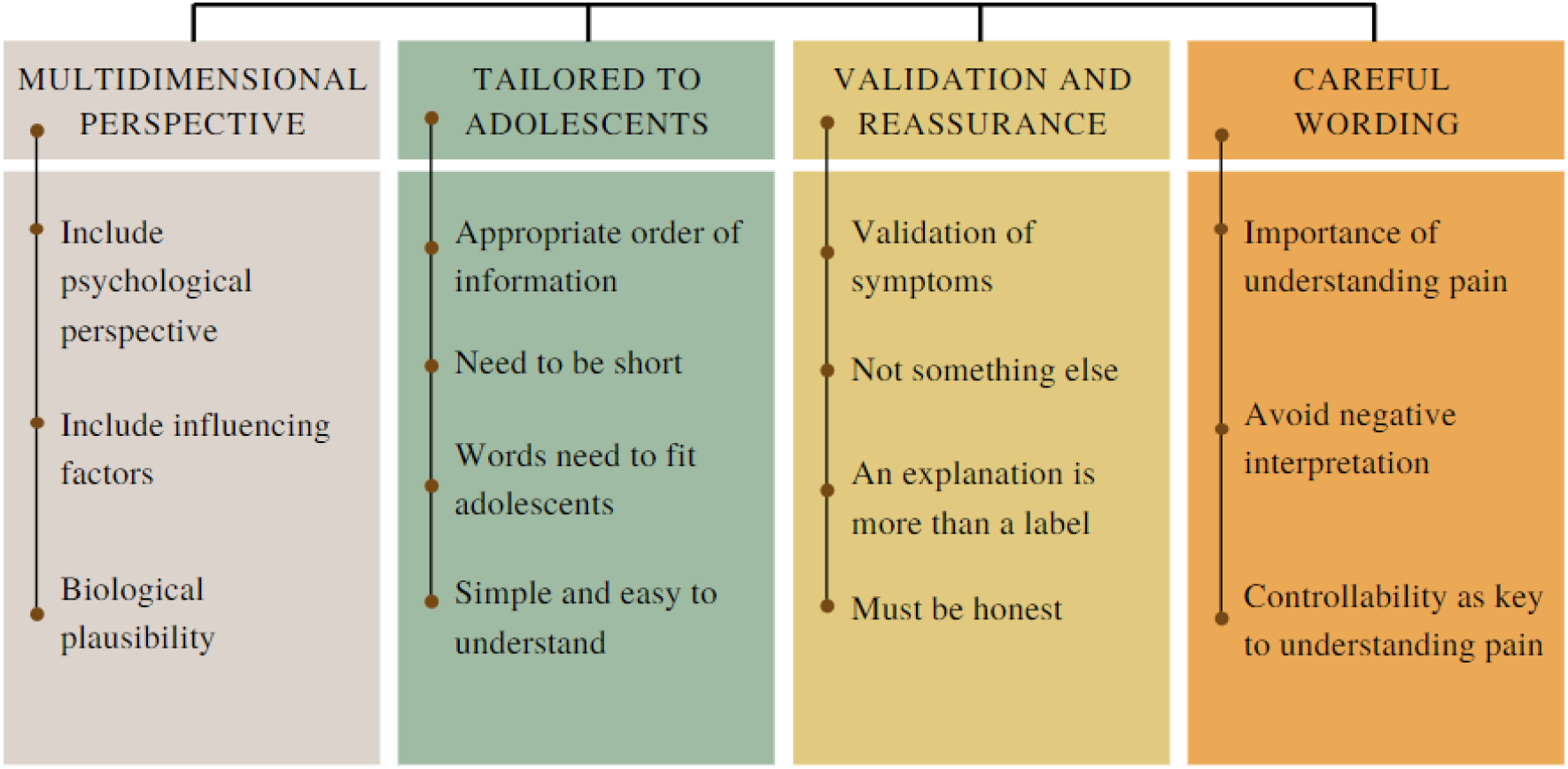
Themes and sub-themes identified from the Argumentative Delphi.

#### 3.2.2. Survey Expert ratings of the credible explanations

According to ratings measured on the Likert scale, we found a positive difference (visual interpretation) in the degree of experts perceived diagnostic uncertainty between Round One and Round Two. Across all three questions, 7 out of 18 experts rated either ‘agree’ or ‘strongly agree’ in Round One and 13 out of 16 experts rated either ‘agree’ or ‘strongly agree’ in Round Two.

### 3.3. Key domains to tailor explanations - Merging themes into domains

We developed three key domains to consider when tailoring credible explanations to adolescents experiencing non-traumatic knee pain. The three key domains were *“What is (diagnosis) and what does it mean?”, “What is causing my knee pain”* and *“How do I manage my knee pain?”* (Figure 3).

**Fig. 3.**
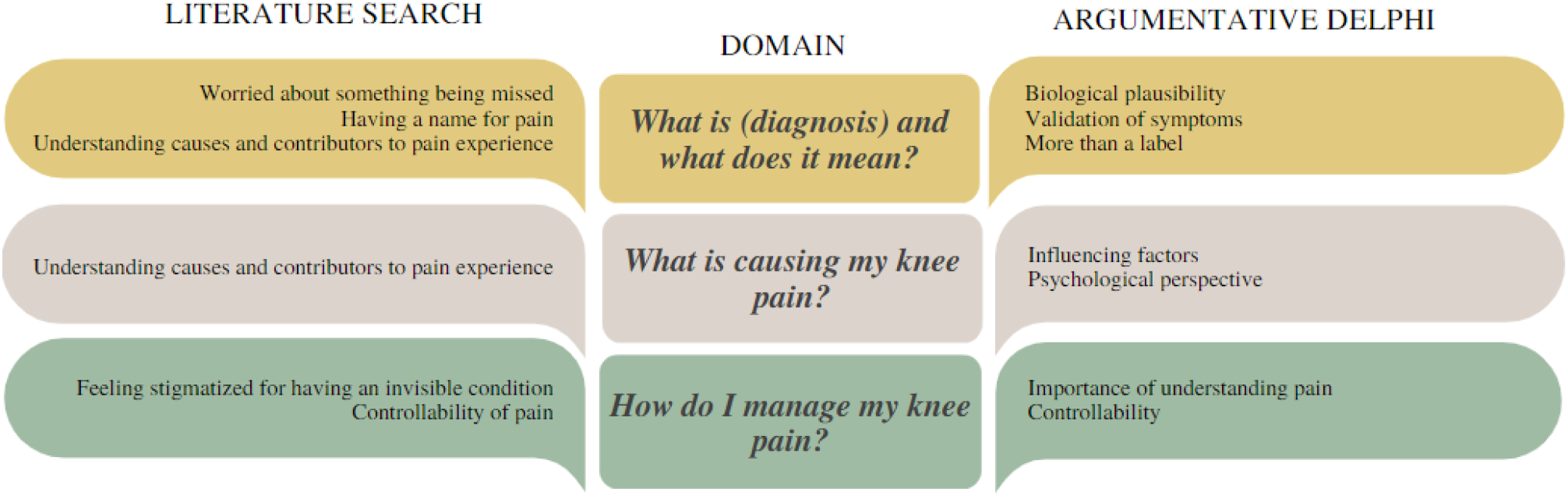
Merging themes from literature search and Argumentative Delphi into domains.

### 3.4. User testing: Think-aloud exercises - adolescents with/without knee pain

We included seven participants (two with knee pain) aged between 8 to 15 years (Table 2). Interview duration ranged between 18 and 45 min (mean duration = 29 min). We included three adolescents for the first session, and four adolescents in Session Two (one with knee pain in each round).

**Table 2.**
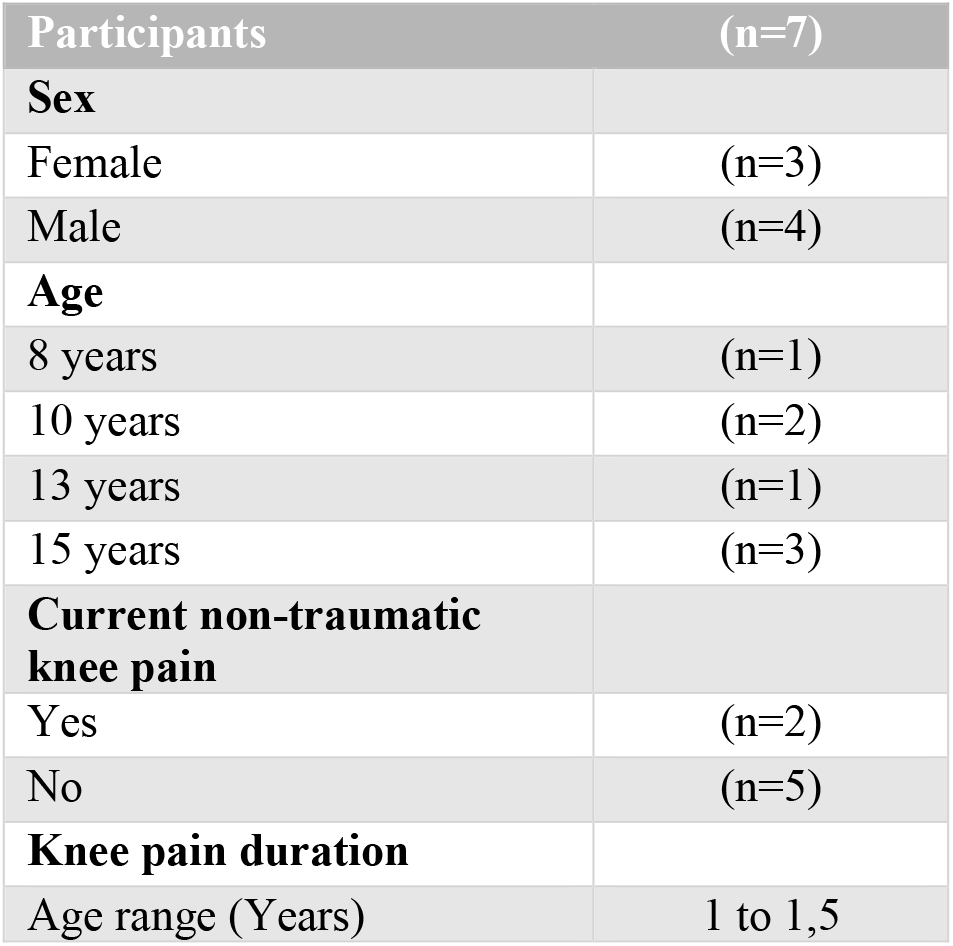
Adolescent demographics: Think-aloud exercises.

The main issues were improper language (medical terms), phrasings that were not understood and long sentences, which allowed ad-hoc corrections [8,27]. Overall, most changes were made in Session One. In Session Two, the reasons for changes were generally the same as in Session One, but the number of comprehension problems was significantly reduced (*Appendix 7)*. During Session Two, most participants expressed great reading flow and changes were at a minimum and required little to no extra mental effort to navigate through the explanations. Participants with current knee pain highlighted the relatable content, such as worrying about pain as important and matched their own lived experience.

### 3.5. Credible explanations for adolescent’s chronic non-traumatic knee pain

We developed six credible explanations for the six most common chronic non-traumatic knee pain diagnoses (Growth pain, Iliotibial Band Syndrome Osgood Schlatter, Patellar Tendinopathy, Patellofemoral Pain, and Sinding-Larsson-Johansson) in English and Danish *(Appendix 8*). The credible explanation for Patellofemoral Pain is visually illustrated in the patient leaflet version (Figure 4).

**Fig. 4.**
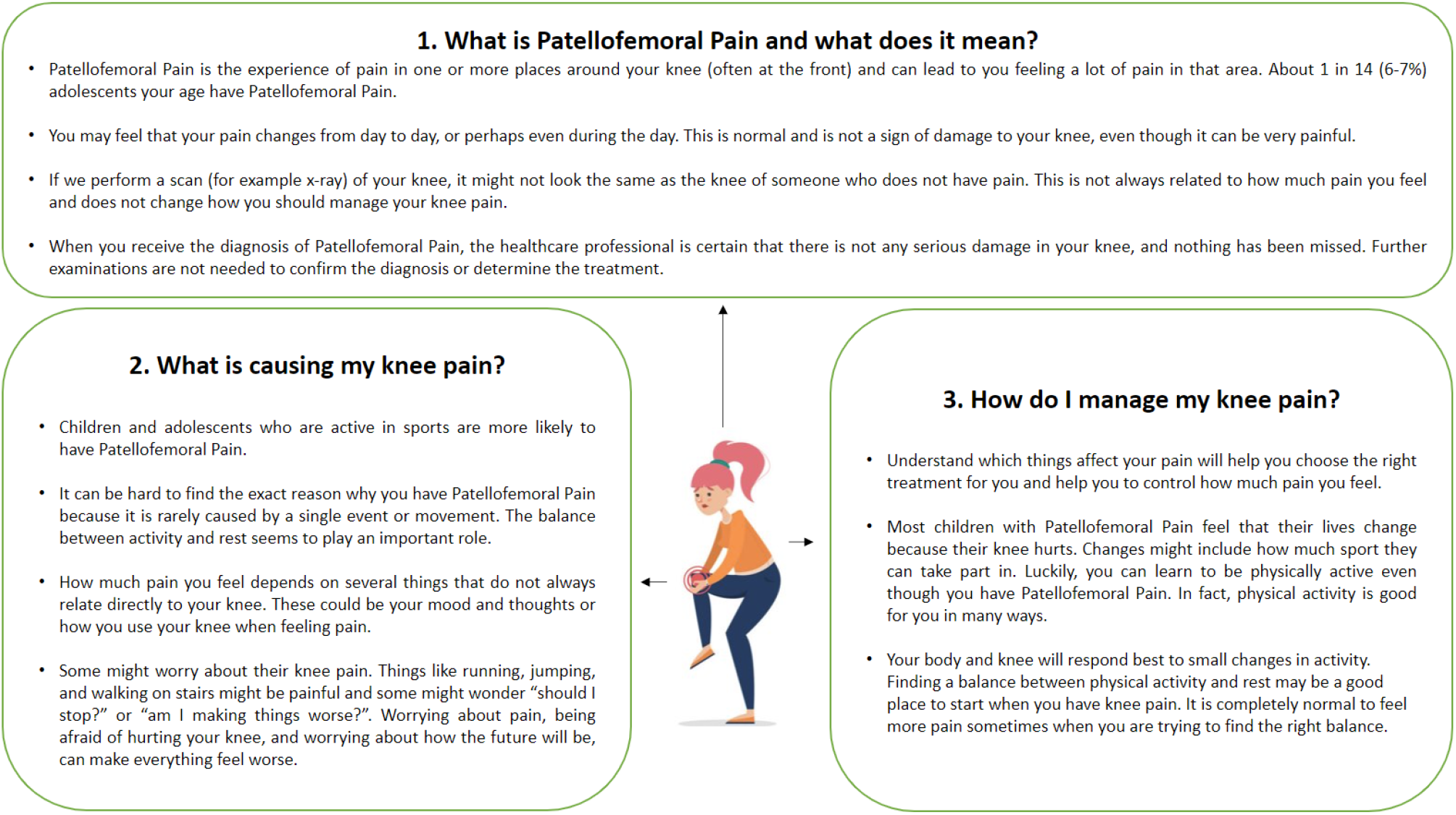
Credible explanation (Patellofemoral Pain). Only Danish versions have been revised based on end-user perspectives (Think-aloud exercises). Therefore, the English version of the credible explanations are our translations exclusively.

## 4. Discussion

This multiple-method study developed credible explanations for the six most common diagnoses of adolescents’ chronic non-traumatic knee pain by integrating perspectives from best research evidence, clinical expertise, and end-users. Through this process, we identified three key domains to consider when tailoring credible explanations for these individuals: *“What is (diagnosis) and what does it mean?”, “What is causing my knee pain”* and *“How do I manage my knee pain?”*. These domains extend the current knowledge and may provide a knowledge-mobilization framework to inform the design for future credible explanations in the context of pediatric CMP.

### 4.1. Explanations of findings

The three key domains all concerned adolescents’ understanding of CMP and ways to manage it, which is consistent with a recent Delphi that explored expert’s opinions on the key messages needed when communicating with young children [65]. Wallwork et al. found that understanding ‘how pain works’, reassurance, normalizing pain, validation and communicating that the child can have some control over the pain are important messages [65]. Their findings highlight the multidimensional nature of CMP, which was also a central theme among our experts. This strengthens the evidence for these domains as the two studies with different methods both identified similar content. Our study further extends previous findings through a deeper understanding of these themes, such as: *“An explanation is more than a label”* or *“Needs to be short”*.

There is an on-going debate about diagnostic labels, their meaning, their clinical value, and their usefulness [18,38,52]. This study suggest that diagnostic labels may have a significant value in the form of external validation; The diagnostic label justified the experience of pain, and even provided societal acceptance, which (sometimes) was a key starting point for acceptance and facilitated the process towards recovery [29,43,46,61]. Our results from the literature search suggest that diagnostic labels are needed, but the associated explanation is important to add meaning to the label and thereby reduce diagnostic uncertainty and feel validated [17,32,46,64].

An underlying trend in themes identified during our literature search was adolescents’ desire for an increased understanding of what pain is. This desire aligns with the goal of *‘making pain understood’* and indicates that illness perceptions are highly influential in the experience and management of adolescent’s CMP [14,24,30]. Khanom et al. found that adolescents’ interpretations and management of pain are directly influenced by how they understand their pain condition [32]. In addition, these findings were supported by Joslin et al.’s finding that understanding the causes and contributors to pain was a key moment that helped move adolescents towards to recovery after treatment for CMP [30]. Results from our study align with previous studies among adolescents and young adults with non-traumatic knee pain suggesting functional theories about knee pain *(i.e*., *individual perception of ‘why do I have pain?’)* are a strong driver of behavior. This highlights that individuals’ negative interpretations can result in counter-productive choices [28,56].

Clinicians’ inability to communicate and explain the cause for pain was a strong theme in the included qualitative studies; Our literature search showed that adolescents express difficulties with the meaning of the diagnosis and what pain is, illustrating the significant opportunity to improve existing patient education strategies [39,46,61]. Clinicians were commonly described to use heterogeneous terminologies and explanations that did not fit with adolescents existing beliefs (i.e., perception of the cause of the pain and expected management) [39,46,60]. Heterogeneous and non-person-centered pain terminologies were highlighted as a core issue for children and adolescents’ that impaired their ability to understand their pain [14,34]. Our findings concerning the role of diagnostic uncertainty in the qualitative studies shed light on a challenging area for clinicians, as it emphasizes the importance of clear communication between clinicians, adolescents, and their parents [34,44,46]. Caution will be required to ensure that CMP explanations do not become more heterogenous as clinicians move from simple biomedical interpretations of pain to a broader and more nuanced biopsychosocial perspective that incorporates pain complexity and reasons for persistence [1,2,64]. Lee et al. found a range of effective and ineffective communication approaches among adolescents suffering from CMP conditions; similar to our study, they emphasized the importance of providing clear and tailored pain management advice for each individual [34]. Together with our results, this highlights the tremendous impact these mixed messages provided by clinicians have; we found adolescents reported their anxiousness was amplified, due to varying explanations and advice from clinicians [39,60]. Through the Delphi process, experts endorsed the importance of validating the pain and paying attention to *psychological perspectives* of pain and *individual contributing factors*, which greatly emphasized CMP as a multidimensional and personal experience.

Research in pediatric chronic pain has increased in recent years; however, studies have requested translation and mobilization of findings into real-world practice [14,34,47]. We aimed to develop credible explanations to be used as patient material and mobilize evidence-based knowledge into practice to improve information consistency and meaning regarding pain terminologies. Our findings highlight the (potential) value of involving end-users when creating patient materials; we made substantial changes based on the think-aloud exercises. This emphasized that clinicians’ language must be adapted to adolescents’ level to ensure correct interpretation of health information [14,15,34]. Reis et al developed a comic book with the purpose of providing validated pain education for children; although their work included involving of end-users, they still found that over a quarter of included children did not feel satisfied with the content and almost a fifth did not feel it helped them understand pain [57]. A plausible explanation for this may be the non-specific nature of many CMP conditions and individual aspects of managing this condition; This may illustrate the subjective differences and preferences clinicians face in everyday practice. The three domains identified, and final explanations are generic and not tailored to every individual’s beliefs. Therefore, they should not be treated as a checklist or one size fits all, but rather as a framework for clinicians to establish common issues related to reducing diagnostic uncertainty, as these domains might capture important information when managing these young individuals. Adolescents with knee pain are not diagnostic categories, but rather individuals experiencing a health challenge, each with their own story, history, and goals, where preferences in communication might be varying according to developmental level and age [15]. Communication (and explanations) should reflect this individuality.

### 4.2. Clinical implications

This study identified key domains that may be addressed to reduce diagnostic uncertainty. Our results clearly indicate that simply naming an adolescent’s pain condition is not the end goal, and clinicians should work to build meaning and understanding around these diagnoses for each person. Based on the qualitative studies, adolescents emphasized that communication skills among clinicians, such as good listening, reassurance and acknowledgement of the pain’s existence have a positive influence on feeling validated [12,30,39,46]. We urge clinicians to ensure terminology is appropriate and consistent to prevent mixed messages; Clinicians may address adolescents’ own perceptions of why they experience pain and explore perceived management for their pain, as this information might provide valuable clinical knowledge for successful rehabilitation.

### 4.3. Future research

Our study may set the first step for a *proof-of-concept method* of knowledge mobilization in the development of credible explanations for painful conditions; this framework will ensure a variety of opinions are included in the developmental process. We hope this can lead to a more systematic and consistent way of developing patient-centered credible explanations of pain conditions in the future. The effectiveness of using our credible explanations should be tested in the future to establish if they lead to superior outcomes among adolescents with non-traumatic knee pain. An important priority for future research is to further explore concepts and domains related to diagnostic uncertainty to enhance understanding of this phenomenon; this may provide promising opportunities to develop new patient education strategies for these young individuals.

### 4.4. Strengths and limitations

Strengths of our study include the engagement with end-users and the employment of a multi-method strategy. Another strength was the development of this iterative process with multiple revisions and adaptations to meet adolescents’ needs. Our study also has certain limitations. First, only one database was used for our systematic searches; relevant papers may not have been included. Second, we included experts with a range of professions and the majority were physiotherapists (n=12). This may bias the opinions and views in the Argumentative Delphi to this profession. Third, a limitation of using an argumentative Delphi is the risk of unclear and unrelated arguments, which made it difficult to interpret the specific reason for the changes [20,63]. Fourth, only one of the six explanations was tested by end-users. Finally, only two participants had knee pain; it cannot be ruled out that some valuable information may have been lost during this process.

## 5. Conclusion

This multiple-method study provided six credible explanations for the six most common diagnoses of chronic non-traumatic knee pain. We identified three essential domains for clinicians to consider when tailoring credible explanations to adolescents experiencing non-traumatic knee pain. These results could inform the design for future credible explanations in the context of pediatric chronic pain.

## Supporting information

Supplementary files - How do we explain painful chronic non-traumatic knee conditions to children and adolescents - A multiple-method study to develop

## Data Availability

All data produced in the present study are available upon reasonable request to the corresponding author.

## Acknowledgements

All authors would like to thank Kristian Damgaard Lyng for helpful discussions during the development of the REDCap survey.

