## Supplementary files - Tables and figures for "How do we explain painful chronic non-traumatic knee conditions to children and adolescents? A multiple-method study to develop credible explanations"

**FIGURE LEGENDS.**

**Fig.1 Multiple-method study design: Iterative process.**

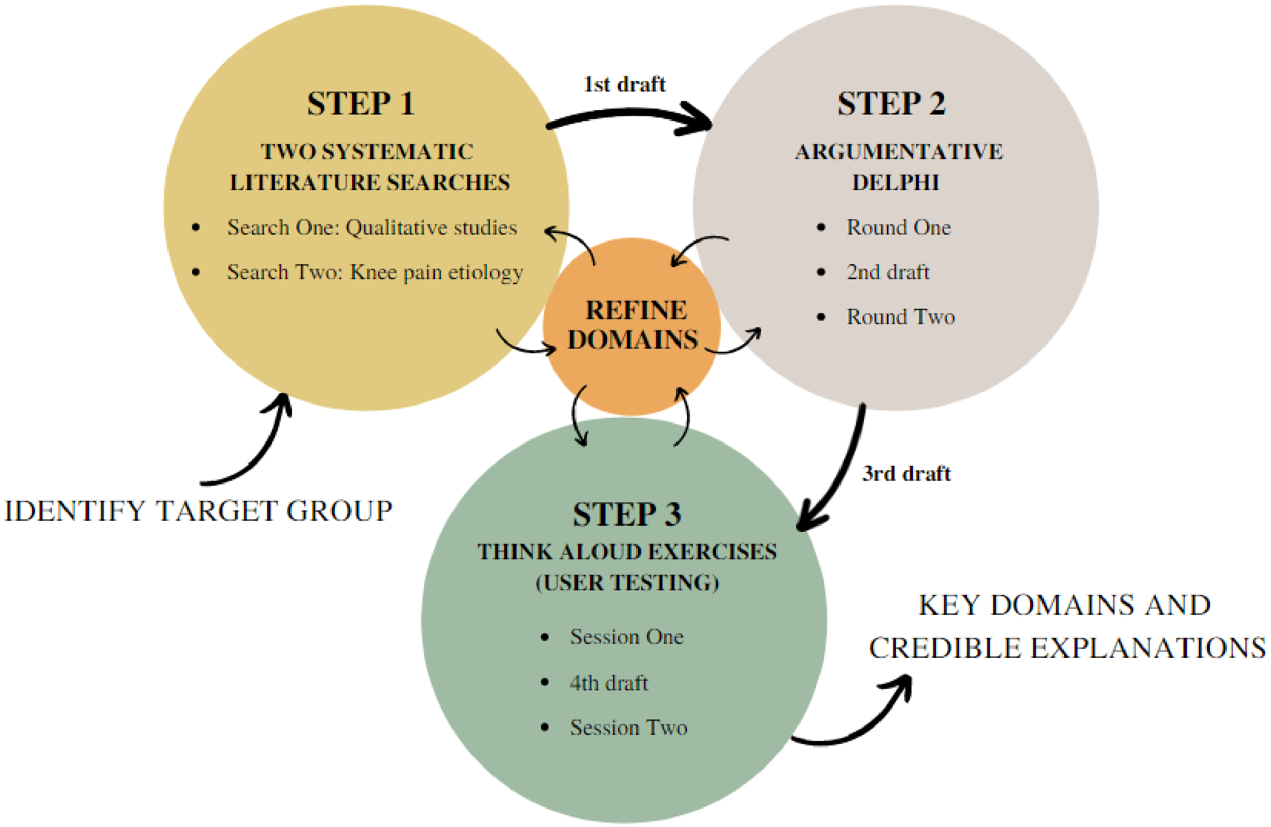

**Fig.2 Themes and sub-themes identified from the Argumentative Delphi.**

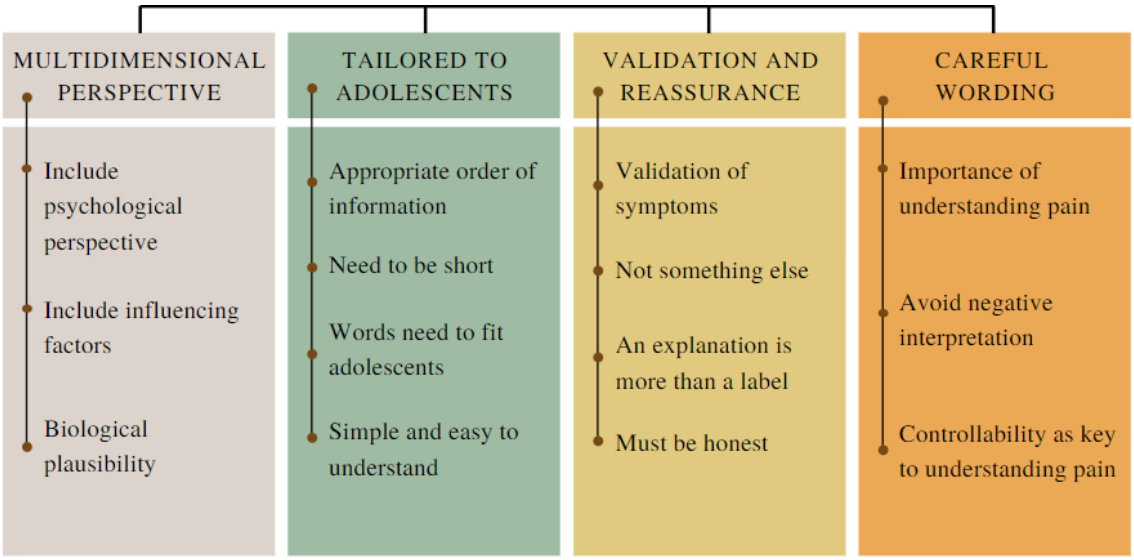

**Fig.3. Merging themes from literature search and Argumentative Delphi into domains.**

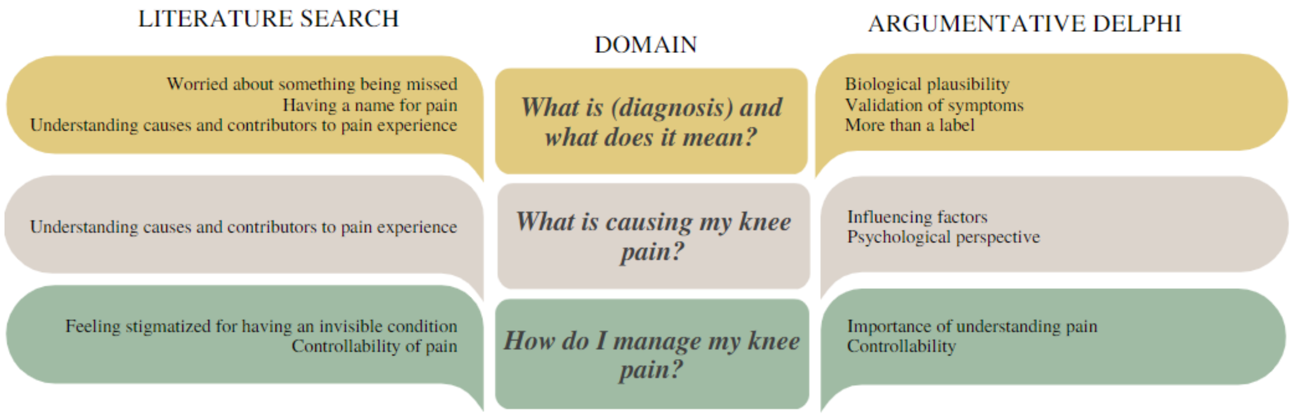

**Fig.4. Credible explanation (Patellofemoral Pain). Only Danish versions have been revised based on end-user perspectives (Think-aloud exercises). Therefore, the English version of the credible explanations are our translations exclusively.**

### 1. What is Patellofemoral Pain and what does it mean?

- Patellofemoral Pain is the experience of pain in one or more places around your knee (often at the front) and can lead to you feeling a lot of pain in that area. About 1 in 14 (6-7%) adolescents your age have Patellofemoral Pain.
- You may feel that your pain changes from day to day, or perhaps even during the day. This is normal and is not a sign of damage to your knee, even though it can be very painful.
- If we perform a scan (for example x-ray) of your knee, it might not look the same as the knee of someone who does not have pain. This is not always related to how much pain you feel and does not change how you should manage your knee pain.
- When you receive the diagnosis of Patellofemoral Pain, the healthcare professional is certain that there is not any serious damage in your knee, and nothing has been missed. Further examinations are not needed to confirm the diagnosis or determine the treatment.

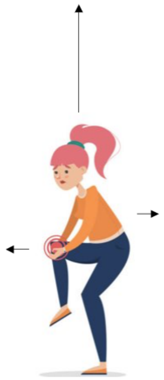

### 2. What is causing my knee pain?

- Children and adolescents who are active in sports are more likely to have Patellofemoral Pain.
- It can be hard to find the exact reason why you have Patellofemoral Pain because it is rarely caused by a single event or movement. The balance between activity and rest seems to play an important role.
- How much pain you feel depends on several things that do not always relate directly to your knee. These could be your mood and thoughts or how you use your knee when feeling pain.
- Some might worry about their knee pain. Things like running, jumping, and walking on stairs might be painful and some might wonder "should I stop?" or "am I making things worse?". Worrying about pain, being afraid of hurting your knee, and worrying about how the future will be, can make everything feel worse.

### 3. How do I manage my knee pain?

- Understand which things affect your pain will help you choose the right treatment for you and help you to control how much pain you feel.
- Most children with Patellofemoral Pain feel that their lives change because their knee hurts. Changes might include how much sport they can take part in. Luckily, you can learn to be physically active even though you have Patellofemoral Pain. In fact, physical activity is good for you in many ways.
- Your body and knee will respond best to small changes in activity. Finding a balance between physical activity and rest may be a good place to start when you have knee pain. It is completely normal to feel more pain sometimes when you are trying to find the right balance.

**TABLES: CAPTIONS AND LEGENDS**

**Table 1. Characteristics of experts: Argumentative Delphi.**

| Experts included | (n=16) |
| --- | --- |
| **Sex** | |
| Male | (n=9) |
| Female | (n=7) |
| **Profession** | |
| Physiotherapist | (n=9) |
| Medical doctor | (n=5) |
| Psychologist | (n=1) |
| Academic researcher | (n=1) |
| **Clinical experience** | |
| 5-10 | (n=3) 21,4% |
| 11-15 | (n=4) 28,6% |
| 15+ | (n=7) 50,0% |
| Missing | (n=2) 12,5% |
| **Working status** | |
| In research/academia and don't see patients on a regular basis. | (n=2) 12,5% |
| In clinical practice seeing patients on a regular basis. | (n=5) 31,3% |
| I share my time between clinical practice and research. | (n=9) 56,3% |
| **Average number of MSK patients/month** | |
| 0 | (n=1) 7,1% |
| 1-5 | (n=3) 21,4% |
| 6-10 | (n=4) 28,6% |
| 11-15 | (n=2) 14,3% |
| 15+ | (n=4) 28,6% |
| **Academic level** | |
| Bachelor’s degree | (n=2) 12,5% |
| Master’s degree | (n=9) 56,25% |
| PhD | (n=4) 25,0% |
| Other | (n=1) 6,3% |
| **Academic research experience** | |
| 0-4 | (n=1) 9,1% |
| 5-10 | (n=3) 27,3% |
| 11-15 | (n=4) 36,4% |
| 15+ | (n=3) 27,3% |
| No academic experience | (n=2) 12,5% |
| **Diagnosis expertise (Multiple options for each expert)** | |

| | |
|---|---|
| Growth Pain | (n=4) |
| Osgood-Schlatter | (n=5) |
| Patellofemoral Pain | (n=7) |
| Patellar Tendinopathy | (n=6) |
| Sinding-Larsen-Johansson | (n=6) |
| Iliotibial Tract Syndrome | (n=1) |

**Table 2. Adolescent demographics: Think-aloud exercises.**

| Participants | (n=7) |
|---|---|
| **Sex** | |
| Female | (n=3) |
| Male | (n=4) |
| **Age** | |
| 8 years | (n=1) |
| 10 years | (n=2) |
| 13 years | (n=1) |
| 15 years | (n=3) |
| **Current non-traumatic knee pain** | |
| Yes | (n=2) |
| No | (n=5) |
| **Knee pain duration** | |
| Age range (Years) | 1 to 1,5 |
