## Supplementary files - How do we explain painful chronic non-traumatic knee conditions to children and adolescents - A multiple-method study to develop for "How do we explain painful chronic non-traumatic knee conditions to children and adolescents? A multiple-method study to develop credible explanations"

### Supplementary Material for: « *How do we explain painful knee conditions to children and adolescents? A multiple-method study to develop credible explanations – Supplementary files.* »

#### Authors
Djurtoft C[1,2,X], Bruun MK[1,X], Riel H[1,2,3] Hoegh MS[2], Darlow B[4], Rathleff MS[1,2]

#### Affiliations
1. Center for General Practice at Aalborg University, Denmark.
2. Department of Health Science and Technology, Faculty of Medicine, Aalborg University, Denmark.
3. Department of Physiotherapy, University College of Northern Denmark, Denmark.
4. Department of Primary Healthcare and General Practice, University of Otago, Wellington, New Zealand.
X. Shared first authorship.

### 1. Appendix 1 – Full description of the two systematic searches

#### 1.1. *Evidence: What information is needed to understand chronic musculoskeletal pain from the perspectives of adolescents and their parents?*

A systematic search was performed to explore needs and experiences from adolescents and their parents' point of view when understanding chronic pain conditions (e.g., chronic pain or non-traumatic knee pain). Eligibility criteria for included papers were qualitative studies *(primary qualitative studies or systematic reviews of qualitative studies)* exploring the experiences related to understanding chronic pain conditions *(i.e., diagnostic uncertainty)* for adolescents (10-19 years old) and/or parents. Only papers in Danish and English were included. The search was performed in MEDLINE (*via PubMed*) in March 2022. The search strategy comprised three key concepts: Diagnostic uncertainty/Chronic pain, adolescents/parents, and qualitative studies. Initially, CD and MKB screened all titles/abstracts independently before deciding which papers to include for full-text reading. We extracted information on participants/setting, aim of study, data collection method (e.g., interview or focus group) and main themes and subthemes. We assessed all themes in the included papers and highlighted key quotations that explored participants' understanding of chronic pain. Themes and subthemes were then extracted by CD and MKB independently and assigned descriptive codes using an inductive process. All codes were grouped into similarity and organized into themes. To allow for building on existing research, we used an index paper to orientate our analysis, as recommended for meta-ethnography [2]. Pincus et al was chosen as our index paper, which includes three key domains for diagnostic uncertainty: *Label/diagnosis, Cause of pain, and Not something else* [15]. If CD or MKB were not able to map a theme or subtheme into the predefined index domains, but the theme added insight, or helped to understand the experience of understanding pain or diagnosis, these themes were organized into additional categories through a collaborative process of constant comparison. The data analysis process was subsequently checked independently by CD and MKB before the final themes (tentative domains) were confirmed by the research team.

##### *1.2.Evidence: What is the etiology for non-traumatic knee pain?*

A systematic literature search was conducted in Medline (via PubMed) in March 2022. Eligibility criteria for included papers were narrative or systematic reviews, describing or investigating the etiology and/or pathogenesis of non-traumatic knee pain. Only papers in Danish and English were included. All patient age groups were included, provided they described etiology and/or pathogenesis of most common non-traumatic knee pain conditions seen in adolescents [5]. Due to risk of outdated information, studies published before 2000 were excluded. The search strategy was developed using medical subject headings and text words related to the six most common knee pain diagnoses as described by Guldhammer et al (Growth pain, Osgood Schlatter, Sinding-Larsson-Johansson, Patellar tendinopathy, Patellofemoral pain, Iliotibial band syndrome); Therefore, five independent searches were performed [5]. The search strategy is available in. We also conducted a hand search including the reference lists of included papers.

Initially, CD and MKB screened all titles/abstracts independently before including papers for full-text reading. After reading the included full texts independently of each other, CD and MKB extracted information from the systematic search relating to *etiology (What is the diagnosis), common causal factors (how does it occur)* and *predisposing factors* independently. After agreements, each condition was described in cooperation by CD and MKB. Conflicting evidence, level of evidence, publishing date and consensus in the literature were considered. If there were any disagreements between CD and MKB, HR and MSR were included for discussions.

##### *1.3.Search strategy: PICo for Qualitative Studies:*

*No filters used.*

| Problem: Diagnostic uncertainty / Chronic pain | Interest: Adolescents | Context: Study type |
|---|---|---|
| "Medically Unexplained Symptoms"[Mesh] OR "Unexplained Symptoms, Medically" OR "Symptoms, Medically Unexplained" OR “Medical unexplained symptoms” OR “Medically unexplained symptoms” or “Diagnostic uncertainty” OR “Diagnostic communication” or | "Paediatric" OR “Pediatric” OR "Child" or "Kids" or "Adolescent" or "Teenager" or "teen" or "Young Athlete" or "Juvenile" or "Paediatrics" or "Immature" or "Adolescents" OR “Parent” OR “Parents” OR “Mother” OR “Father” | “Qualitative” OR “Interview*” OR “experienc*” OR “perspective*” OR “understanding*” OR “perceive” OR “perceives” OR “semi-structured” OR “perceiving” |

| | | |
|---|---|---|
| "credible explanation" OR "Diagnostic certainty" OR "musculoskeletal pain" OR "Longstanding pain" OR "Chronic pain" OR "persistent pain" | | |
| Hits: 69,651 | Hits: 4,188,428 | Hits: 2,967,646 |
| Hits: 3,238<br><br>(("musculoskeletal pain" OR "Longstanding pain" OR "Chronic pain" OR "persistent pain" OR "Medically Unexplained Symptoms"[Mesh] OR "Unexplained Symptoms, Medically" OR "Symptoms, Medically Unexplained" OR "Medical unexplained symptoms" OR "Medically unexplained symptoms" or "Diagnostic uncertainty" OR "Diagnostic communication" or "credible explanation" OR "Diagnostic certainty") AND ("Paediatric" OR "Pediatric" OR "Child" or "Kids" or "Adolescent" or "Teenager" or "teen" or "Young Athlete" or "Juvenile" or "Paediatrics" or "Immature" or "Adolescents" OR "Parent" OR "Parents" OR "Mother" OR "Father")) AND ("Qualitative" OR "Interview*" OR "experienc*" OR "perspective*" OR "understanding*" OR "perceive" OR "perceives" OR "semi-structured" OR "perceiving") | | |

#### *1.4.Search strategy: Knee pain etiology:*

*No filters used.*

| Knee pain/injuries | Children/adolescents | Aetiology/treatment | Review |
|---|---|---|---|
| **Growth pain** | | | |
| **PubMed: 17,789** | 3,704,344 hits | 4,064,375 hits | 3,861,578 hits |
| **Combined: 250** | (Knee pain + Aetiology + Children):<br>**1,218 hits** | (Knee pain + Aetiology):<br>**6,628 hits** | (Knee pain + Aetiology + Review):<br>**1,589 hits** |
| "Growth pain*" or "growing pain*" or "growing-pain*" or "leg pain" or "musculoskeletal pain*" or "benign nocturnal limb pain*" or "lower extremity pain*" | "Child" or "Kids" or "Adolescent" or "Teenager" or "teen" or "Young Athlete" or "Juvenile" or "Junior Paediatric" or "Immature" | "etiology" or "aetiology" or "cause" or "causes" or "causation" or "pathogenesis" or "pathophysiology" or "pathology" or "aetiopathogenesis" | "Syntheses" OR "metaanaly*" OR "meta-analy*" OR "review" OR "systematic-review" or "synthesize" |
| **Osgood schlatter** | | | |
| **(PubMed: 8,073)** | 3,704,344 hits | 4,064,375 hits | 3,861,578 hits |
| **Combined: 218** | (Knee pain + Aetiology + Children):<br>**1,093 hits** | (Knee pain + Aetiology):<br>**3,242 hits** | (Knee pain + Aetiology + Review):<br>**562 hits** |
| "Osteochondrosis"[Mesh] or "Osteochondrosis" OR "Patellar Ligament"[Mesh] or "OSD" "Patellar Ligament" OR "Growth Plate"[Mesh] or "Growth Plate" OR "osgood schlatter*" OR "patellar ligament" | "Child" or "Kids" or "Adolescent" or "Teenager" or "teen" or "Young Athlete" or "Juvenile" or "Junior Paediatric" or "Immature" | "etiology" or "aetiology" or "cause" or "causes" or "causation" or "pathogenesis" or "pathophysiology" or "pathology" or "aetiopathogenesis" | "Syntheses" OR "metaanaly*" OR "meta-analy*" OR "review" OR "systematic-review" or "synthesize" |

| | | | |
|---|---|---|---|
| **Patellar tendinopathy** | | | |
| **(PubMed: 5,005)** | 3,704,344 hits | 4,064,375 hits | 3,861,578 hits |
| **Combined: 44** | (Knee pain + Aetiology + Children):<br>**353 hits** | (Knee pain + Aetiology):<br>**1,478 hits** | (Knee pain + Aetiology + Review):<br>**251 hits** |
| "Patella tendon*" or "patellar tendon*" or "Knee tendin*" or "patella tendin*" or "patellar tendin*" or "jumpers knee" or "jumper's knee" or "Sinding-Larsen-Johansson" | "Child" or "Kids" or "Adolescent" or "Teenager" or "teen" or "Young Athlete" or "Juvenile" or "Junior Paediatric" or "Immature" | "etiology" or "aetiology" or "cause" or "causes" or "causation" or "pathogenesis" or "pathophysiology" or "pathology" or "aetiopathogenesis" | "Syntheses" OR "metaanaly*" OR "meta-analy*" OR "review" OR "systematic-review" or "synthesize" |
| **Patellofemoral pain** | | | |
| **(PubMed: 7,939)** | 3,704,344 hits | 4,064,375 hits | 3,861,578 hits |
| **Combined: 96** | (Knee pain + Aetiology + Children):<br>**572 hits** | (Knee pain + Aetiology):<br>**2,208 hits** | (Knee pain + Aetiology + Review):<br>**474 hits** |
| "Patellofemoral Pain Syndrome"[Mesh] or "Patellofemoral Pain Syndrome" or "Knee pain" or "PFP" or "chondromalacia" or "patell*" OR "femoropatell*" OR "anterior knee*" OR "peripatell*" "kneecap" OR "patellofemoral" OR "patello-femoral" | "Child" or "Kids" or "Adolescent" or "Teenager" or "teen" or "Young Athlete" or "Juvenile" or "Junior Paediatric" or "Immature" | "etiology" or "aetiology" or "cause" or "causes" or "causation" or "pathogenesis" or "pathophysiology" or "pathology" or "aetiopathogenesis" | "Syntheses" OR "metaanaly*" OR "meta-analy*" OR "review" OR "systematic-review" or "synthesize" |
| **Iliotibial band syndrome** | | | |
| **PubMed: (902)** | 3,704,344 hits | 4,064,375 hits | 3,861,578 hits |
| **Combined: 7** | (Knee pain + Aetiology + Children):<br>**41 hits** | (Knee pain + Aetiology):<br>**262 hits** | (Knee pain + Aetiology + Review):<br>**58 hits** |
| "Iliotibial band syndrome" or "ITBS" or "Iliotibial tract" or "Maissiat's band" or "iliotibial band friction syndrome" or "iliotibial band strain" | "Child" or "Kids" or "Adolescent" or "Teenager" or "teen" or "Young Athlete" or "Juvenile" or "Junior Paediatric" or "Immature" | "etiology" or "aetiology" or "cause" or "causes" or "causation" or "pathogenesis" or "pathophysiology" or "pathology" or "aetiopathogenesis" | "Syntheses" OR "metaanaly*" OR "meta-analy*" OR "review" OR "systematic-review" or "synthesize" |

#### 2. Appendix 2 – Flow-chart: Delphi process

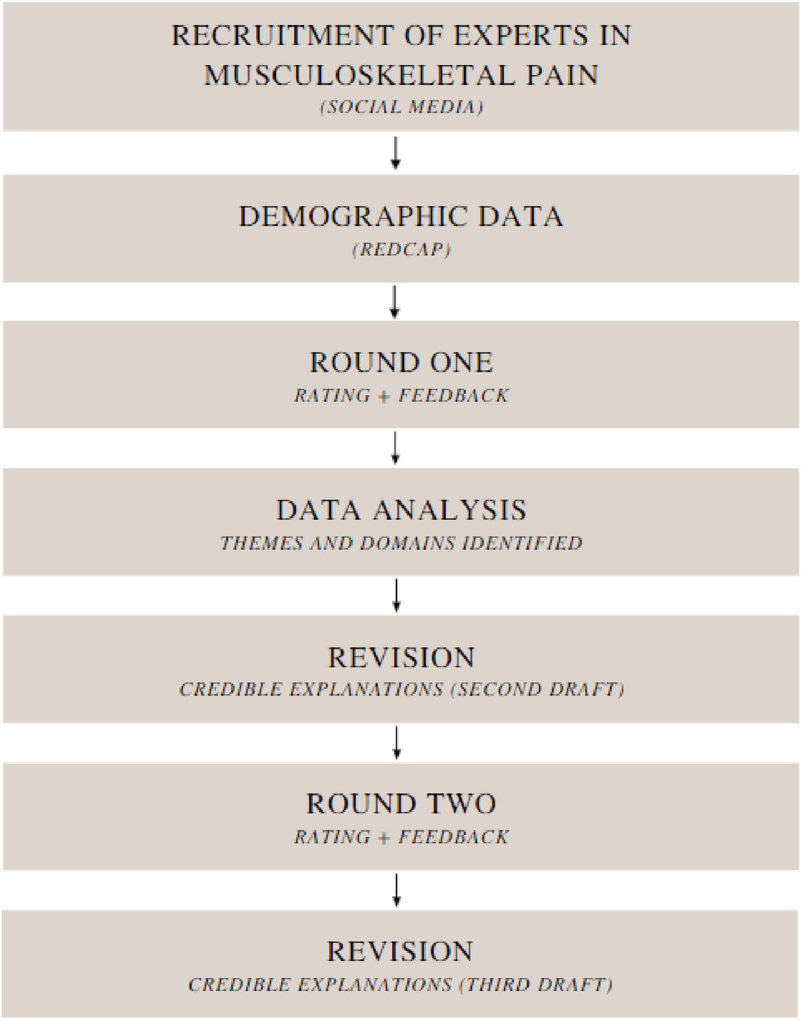

Figure 1. Argumentative Delphi (Flow chart)

#### 3. Appendix 3 – Expert Call-out

ARE YOU INTERESTED IN IMPROVING CLINICAL PRACTICE FOR MANAGING ADOLESCENT KNEE PAIN?

Researchers from Aalborg University and Center for General Practice are conducting a project, where the aim is to improve current management of adolescent non-traumatic knee pain. We want to ensure that kids and adolescents receive the best possible explanation for their knee pain by means of a leaflet, which will be developed by the research team based on best available knowledge.

We are looking for healthcare professionals in musculoskeletal pain with at least 5 years of experience and/or currently working with research in this particular field. Your expertise will contribute to shaping how future healthcare professionals will explain a painful experience to adolescents with non-traumatic knee pain.

#### 4. Appendix 4 – Likert (Pincus et al)

| | | | | |
|---|---|---|---|---|
| **1. The description provides a clear explanation of the label/diagnosis** | | | | |
| Strongly disagree | Disagree | Neither agree nor disagree | Agree | Strongly agree |
| **2. The description provides a clear explanation about why the patient may experience knee pain.** | | | | |
| Strongly disagree | Disagree | Neither agree nor disagree | Agree | Strongly agree |
| **3. The description provides a clear explanation regarding no other diagnosis is indicated.** | | | | |
| Strongly disagree | Disagree | Neither agree nor disagree | Agree | Strongly agree |

#### 5. Appendix 5 – Summary of systematic searches

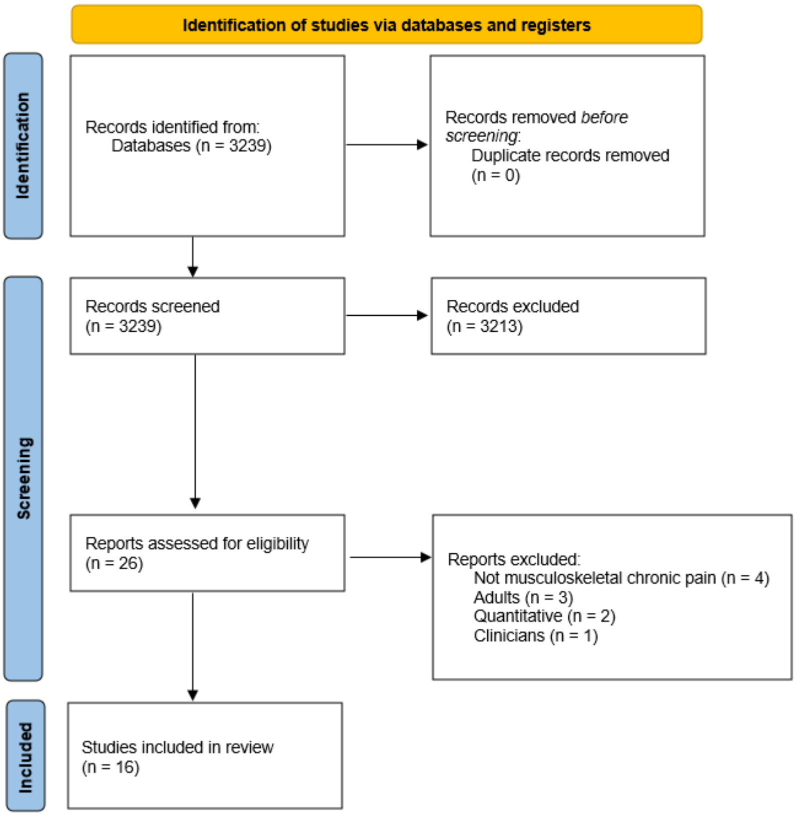

*Figure - Flow-chart: Systematic search of qualitative studies.*

*Table - Summary of literature searches for knee pain etiology.*

| Systematic search | Records screened (n=) | Hand search (n=) | Full text (n=) | Studies included (n=) |
|---|---|---|---|---|
| Growing pain | 1.589 | 1 | 21 | 8 |
| Osgood-Schlatter disease | 562 | 1 | 17 | 8 |
| Tendinopathy + Sinding-Larsson-Johansson | 251 | 3 | 29 | 17 |
| Patellofemoral Pain | 474 | 0 | 33 | 23 |
| Iliotibial band syndrome | 58 | 0 | 12 | 8 |
| **Total** | **2.934** | **5** | **112** | **64** |

#### 6. Appendix 6 – Description of the five themes from qualitative search.

##### *(1) **Understanding causes and contributors to pain experience.***

Adolescents' need for understanding causes and contributors to pain was explored in twelve studies[3,4,6–10,12–14,16,18]. Adolescents felt frustrated when clinicians were unable to explain or understand their pain experiences. Participants described that having a name for their pain (i.e., diagnosis) was not enough to ensure diagnostic certainty, because they needed further information beyond the label for why they were in pain, such as what factors causing/contributing to their experience. Many participants expressed difficulties with the meaning of their diagnosis, because they did not always understand the explanations provided by their clinicians, as they did not align with existing beliefs. This lack of understanding for causes and contributors limited their ability to stay active and engage in recommended therapy. Participants expressed interest in gaining knowledge about pain and understanding causes and contributors for their experience to learn how to prevent incidence of pain and explain the condition to their peers. One study described the understanding of causes and contributors to pain as *"a perfect storm"* and *"a pivotal moment"* in rehabilitation towards optimism and belief that they could get better [8].

##### *(2) **Feeling stigmatized for having an invisible condition.***

Participants in nine studies felt stigmatized for having an invisible condition and highlighted the need for validation of their symptoms and reassurance [3,4,7,9–11,14,16,18]. Invisibility of chronic pain was the most frequently described source of pain-related stigma, contributing to diagnostic uncertainty and lack of understanding and support from others.  The absence of visible signs of pain brought up doubts regarding the symptoms' truthfulness and the constant change of adolescent's attitude towards pain resulted in stigmatization and lack of validation or a deepening sense of isolation. The invisibility of chronic pain often resulted in adolescents felt disbelieved or dismissed by others, including parents, teachers, peers, and clinicians. This resulted in pain-related stigma, accusing adolescents of faking, or exaggerating their pain for attention and further complicated their interactions with health-care professionals. Adolescents raised the importance of reassurance and acknowledgement of their pain experience as an important part of feeling validated.

***(3) <u>Having a name for pain.</u>***

Having a name for pain was identified in eight studies [1,7,11,12,14,16–18]. Participants predominantly reported values of relief, enlightenment, and societal acceptance to the existence of a diagnosis. The absence of diagnosis represented a significant barrier to their ability to live well and remain positive and contributing to perceived lack of understanding by family, friends, and school. Some participants felt angry and distressed towards the healthcare system and viewed is as incompetent, due to the clinicians failed attempt to diagnose their condition. This resulted in experiences of being abandoned. Having a name for their pain helped participants to find strategies and solutions to manage their symptoms, would relieve them from feeling lonely. Both parents and adolescents described a diagnosis as external validation that justified the experience of pain. The 'right' diagnosis had to fit their beliefs of their pain to truly accept it, as the name was used as justification for the pain.

***(4) <u>Controllability of pain.</u>***

The participants' experience of pain and its controllability emerged as a theme in six studies [6,8,9,13,17,18]. Controllability of pain refers to the lack of control some adolescents experience during periods of diagnostic uncertainty. This was not the case for all adolescents, as some believed that they were completely in control of their situation and could manage their own recovery. Adolescents described frustration with diagnostic uncertainty and some mentioned being *'defeated'* by their pain, which led to loss of control over their life and lack of support from social environment. The unpredictability of pain disrupted their hope for the future. Participants wanted to gain control of pain and learn effective management strategies to live normally with pain.

***(5) <u>Worried by something missing.</u>***

Participants highlighted the need for diagnostic certainty in terms of ruling out other diagnoses and was consistent in four studies [1,10,12,14]. Participants outlined the impact of getting the 'right' diagnosis and the fact that nothing has been missed, was a positive step towards accept and successful management. Participants reported a mistrust in the medical system and doubted its competencies, when clinicians were unable to provide satisfactory explanation that reduced diagnostic uncertainty. Participants believing that something serious was happening was common and negative test results did not provide diagnostic certainty or relief. Diagnostic uncertainty caused both adolescents and parents to worry that something serious might have been missed and the absence of solutions could lead them to try alternative solutions, such as complementary medicines or pharmacological treatment. Consequently, this led to frustration and parents reported that they kept searching for the ‘right’ diagnosis until they found a clinician who would provide an acceptable answer.

#### 7. Appendix 7 – Think aloud changes

- Round 1
  - Suggested changes:
    - Easier language
    - Remove long and medical words
  - Positive feedback
    - Relatable content regarding worrying about pain
- Round 2
  - Suggested changes
    - Needs to be short and informative
    - Adding more specific pain location (e.g., bump at tibial tuberosity)
    - Remove long and medical words
    - Examples of what types of sports might cause (diagnosis)

#### 8. Appendix 8 - Credible explanation – Final draft for six diagnoses (English versions only).

Only Danish versions have been revised based on end-users (Think-aloud exercises). The English version of the credible explanations are our translations exclusively. The Danish version was made to fit into a context with medical doctors. Contact the corresponding author to request access to Danish versions.

##### 8.1.English

##### *1. Growth Pain (English)*

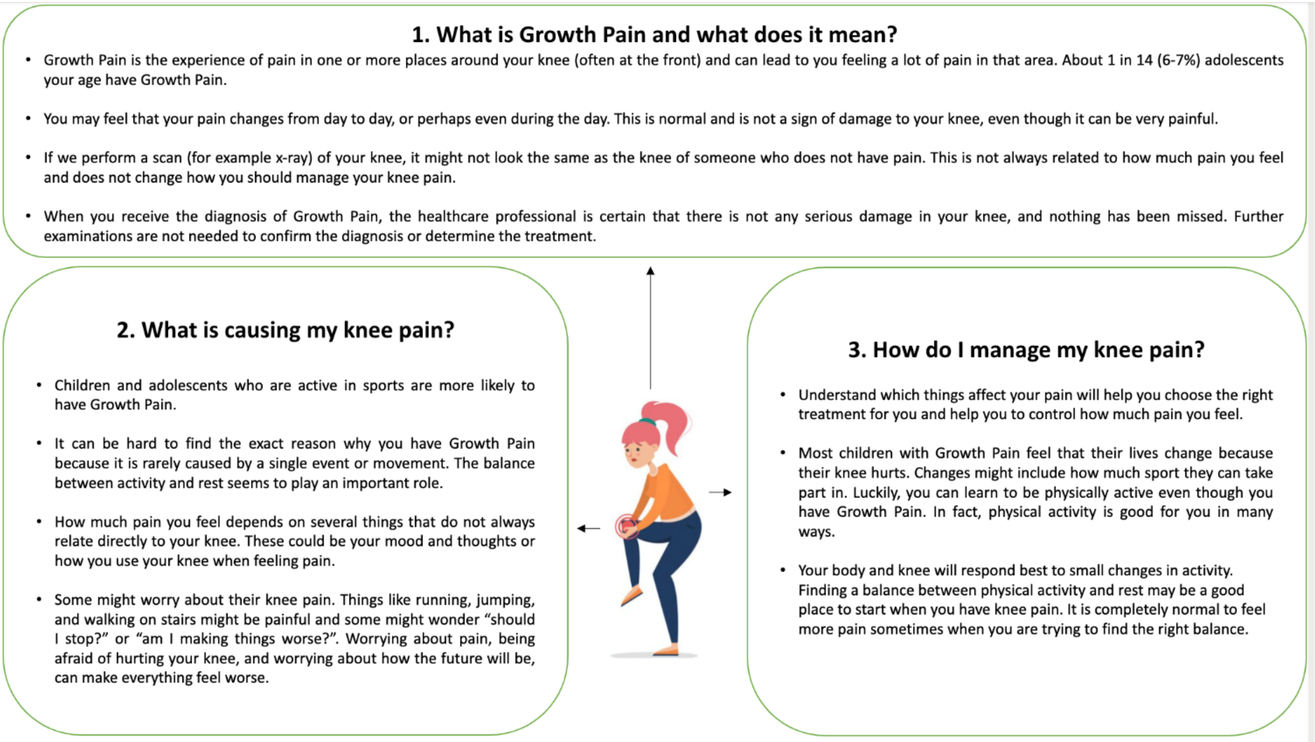

**What is Growth Pain and what does it mean?**

Growth Pain is the experience of pain in one or more places around your knee (often at the front), and can lead to you feeling a lot of pain in that area. About 1 in 14 (6-7%) adolescents your age have Growth Pain.

You may feel that your pain changes from day to day, or perhaps even during the day. This is normal and is not a sign of damage to your knee, even though it can be very painful.

If we perform a scan (for example x-ray) of your knee, it might not look the same as the knee of someone who does not have pain. This is not always related to how much pain you feel and does not change how you should manage your knee pain.

When you receive the diagnosis of Growth Pain, the healthcare professional is certain that there is not any serious damage in your knee, and nothing has been missed. Further examinations are not needed to confirm the diagnosis or determine the treatment.

**What is causing my knee pain?**

Children and adolescents who are active in sports are more likely to have Growth Pain.

It can be hard to find the exact reason why you have Growth Pain because it is rarely caused by a single event or movement. The balance between activity and rest seems to play an important role.

How much pain you feel depends on several things that do not always relate directly to your knee. These could be your mood and thoughts or how you use your knee when feeling pain.

Some might worry about their knee pain. Things like running, jumping, and walking on stairs might be painful and some might wonder “should I stop?” or “am I making things worse?”. Worrying about pain, being afraid of hurting your knee, and worrying about how the future will be, can make everything feel worse.

**How do I manage my knee pain?**

Understand which things affect your pain will help you choose the right treatment for you and help you to control how much pain you feel.

Most children with Growth Pain feel that their lives change because their knee hurts. Changes might include how much sport they can take part in. Luckily, you can learn to be physically active even though you have Growth Pain. In fact, physical activity is good for you in many ways.

Your body and knee will respond best to small changes in activity. Finding a balance between physical activity and rest may be a good place to start when you have knee pain. It is completely normal to feel more pain sometimes when you are trying to find the right balance.

#### 2. *Iliotibial Band Syndrome (English)*

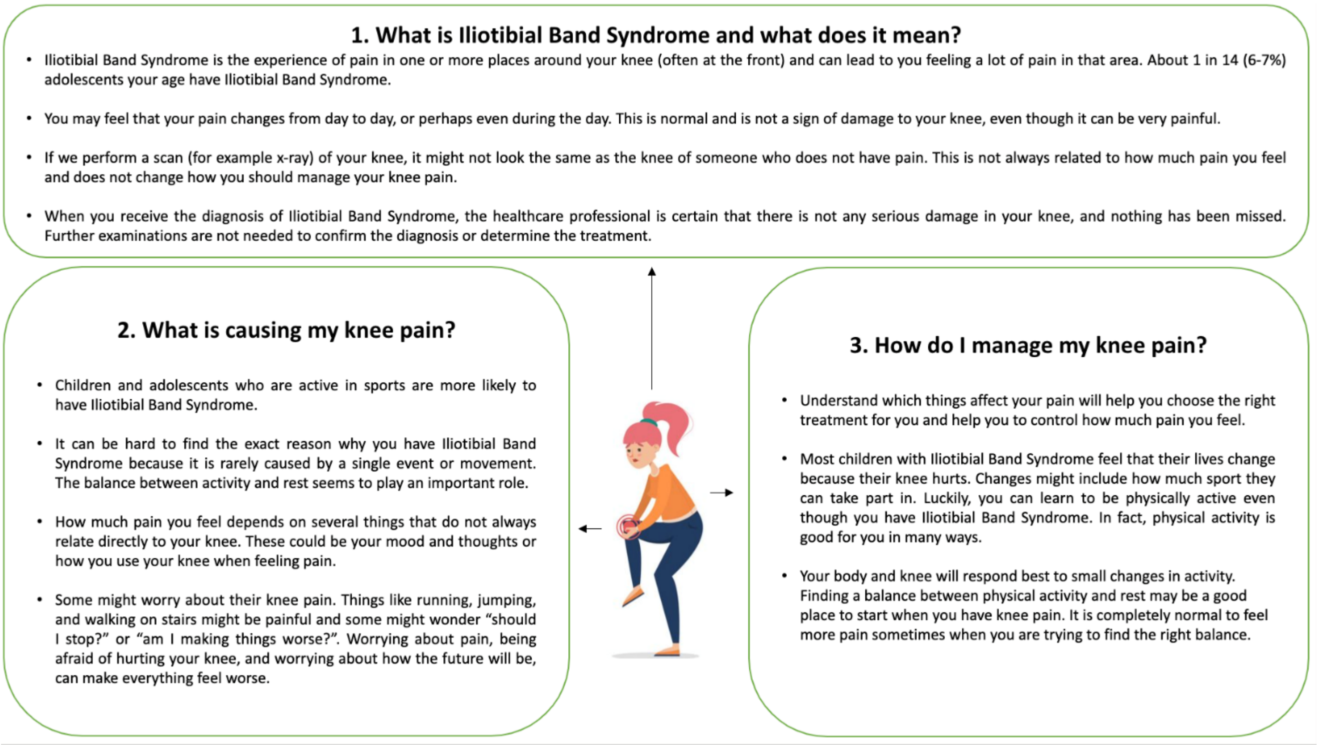

**What is Iliotibial Band Syndrome and what does it mean?**

Iliotibial Band Syndrome is the experience of pain in one or more places around your knee (often at the front), and can lead to you feeling a lot of pain in that area. About 1 in 14 (6-7%) adolescents your age have Iliotibial Band Syndrome.

You may feel that your pain changes from day to day, or perhaps even during the day. This is normal and is not a sign of damage to your knee, even though it can be very painful.

If we perform a scan (for example x-ray) of your knee, it might not look the same as the knee of someone who does not have pain. This is not always related to how much pain you feel and does not change how you should manage your knee pain.

When you receive the diagnosis of Iliotibial Band Syndrome, the healthcare professional is certain that there is not any serious damage in your knee, and nothing has been missed. Further examinations are not needed to confirm the diagnosis or determine the treatment.

**What is causing my knee pain?**

Children and adolescents who are active in sports are more likely to have Iliotibial Band Syndrome.

It can be hard to find the exact reason why you have Iliotibial Band Syndrome because it is rarely caused by a single event or movement. The balance between activity and rest seems to play an important role.

How much pain you feel depends on several things that do not always relate directly to your knee. These could be your mood and thoughts or how you use your knee when feeling pain.

Some might worry about their knee pain. Things like running, jumping, and walking on stairs might be painful and some might wonder “should I stop?” or “am I making things worse?”. Worrying about pain, being afraid of hurting your knee, and worrying about how the future will be, can make everything feel worse.

**How do I manage my knee pain?**

Understand which things affect your pain will help you choose the right treatment for you and help you to control how much pain you feel.

Most children with Iliotibial Band Syndrome feel that their lives change because their knee hurts. Changes might include how much sport they can take part in. Luckily, you can learn to be physically active even though you have Iliotibial Band Syndrome. In fact, physical activity is good for you in many ways.

Your body and knee will respond best to small changes in activity. Finding a balance between physical activity and rest may be a good place to start when you have knee pain. It is completely normal to feel more pain sometimes when you are trying to find the right balance.

#### 3. *Osgood Schlatter Disease (English)*

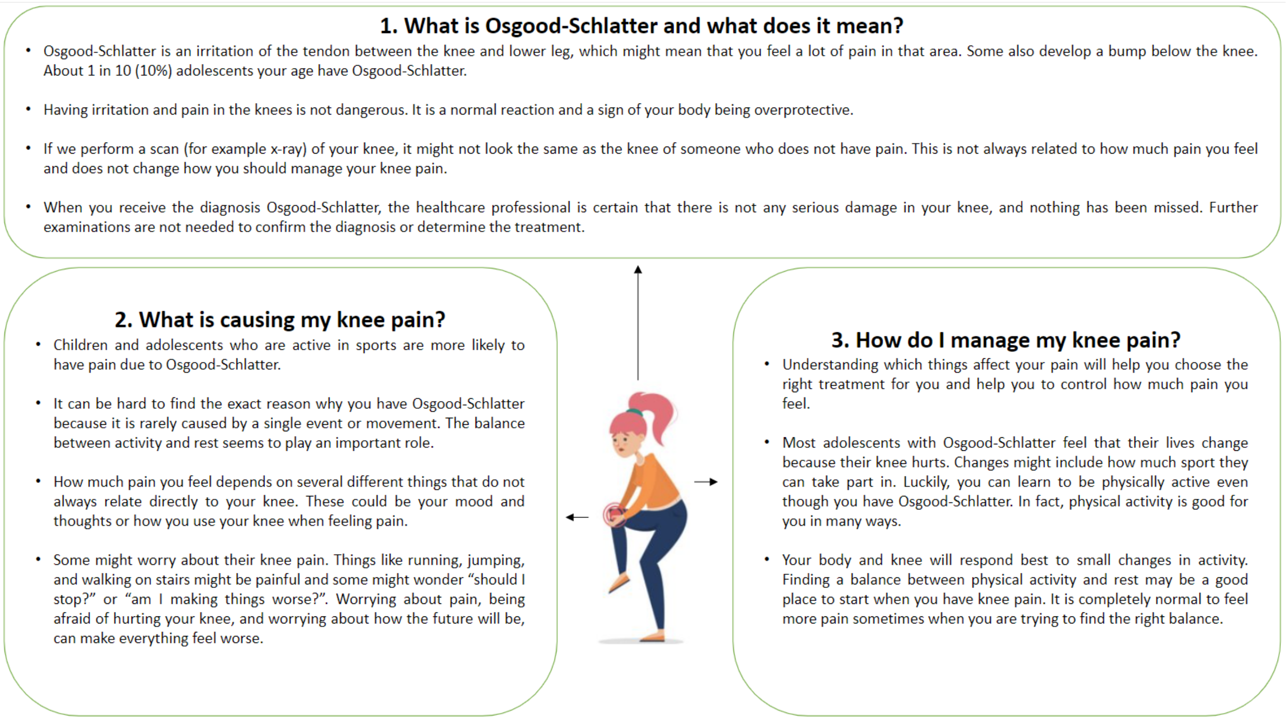

**What is Osgood-Schlatter and what does it mean?**

Osgood-Schlatter is an irritation of the tendon between the knee and lower leg, which might mean that you feel a lot of pain in that area. Some also develop a bump below the knee. About 1 in 10 (10%) adolescents your age have Osgood-Schlatter.

Having irritation and pain in the knees is not dangerous. It is a normal reaction and a sign of your body being overprotective.

If we perform a scan (for example x-ray) of your knee, it might not look the same as the knee of someone who does not have pain. This is not always related to how much pain you feel and does not change how you should manage your knee pain.

When you receive the diagnosis Osgood-Schlatter, the healthcare professional is certain that there is not any serious damage in your knee, and nothing has been missed. Further examinations are not needed to confirm the diagnosis or determine the treatment.

**What is causing my knee pain?**

Children and adolescents who are active in sports are more likely to have pain due to Osgood-Schlatter.

It can be hard to find the exact reason why you have Osgood-Schlatter because it is rarely caused by a single event or movement. The balance between activity and rest seems to play an important role.

How much pain you feel depends on several different things that do not always relate directly to your knee. These could be your mood and thoughts or how you use your knee when feeling pain.

Some might worry about their knee pain. Things like running, jumping, and walking on stairs might be painful and some might wonder “should I stop?” or “am I making things worse?”. Worrying about pain, being afraid of hurting your knee, and worrying about how the future will be, can make everything feel worse.

**How do I manage my knee pain?**

Understanding which things affect your pain will help you choose the right treatment for you and help you to control how much pain you feel.

Most adolescents with Osgood-Schlatter feel that their lives change because their knee hurts. Changes might include how much sport they can take part in. Luckily, you can learn to be physically active even though you have Osgood-Schlatter. In fact, physical activity is good for you in many ways.

Your body and knee will respond best to small changes in activity. Finding a balance between physical activity and rest may be a good place to start when you have knee pain. It is completely normal to feel more pain sometimes when you are trying to find the right balance.

#### *4. Patellar Tendinopathy (English)*

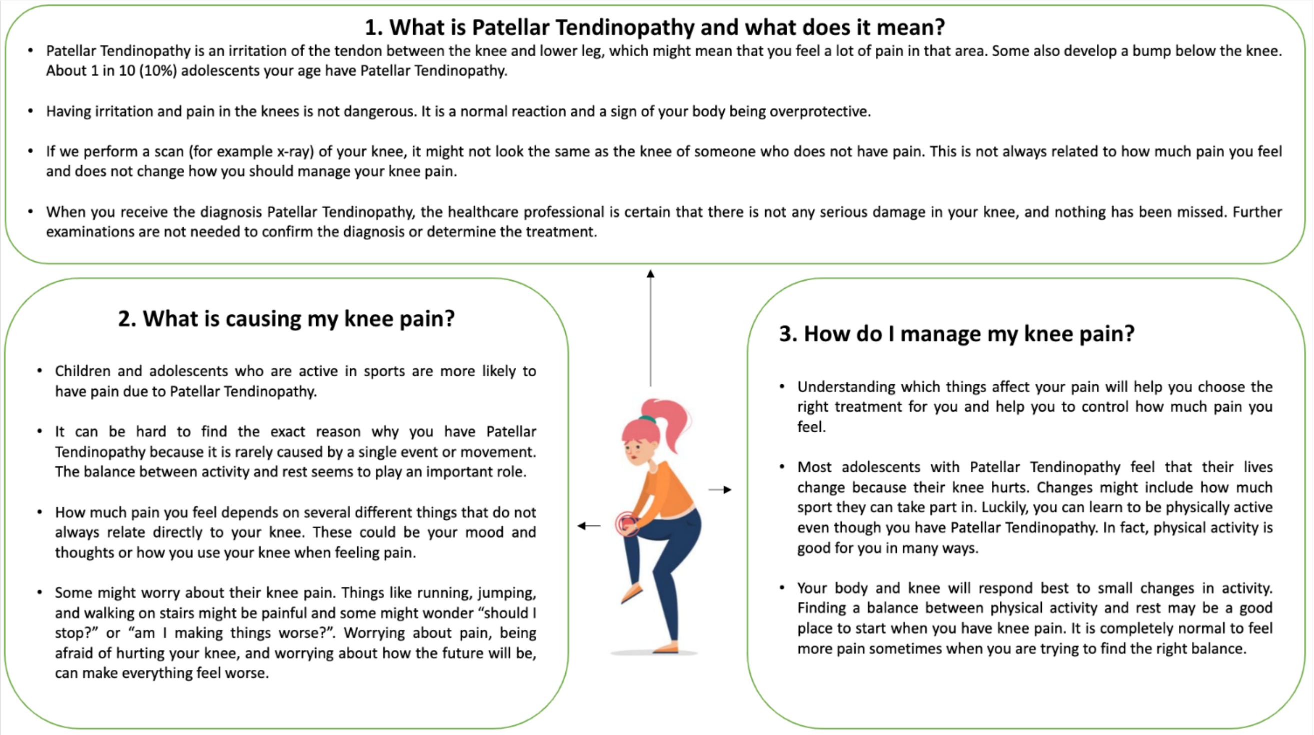

**What is Patellar Tendinopathy and what does it mean?**

Patellar Tendinopathy is an irritation of the tendon between the knee and lower leg, which might mean that you feel a lot of pain in that area. Some also develop a bump below the knee. About 1 in 10 (10%) adolescents your age have Patellar Tendinopathy.

Having irritation and pain in the knees is not dangerous. It is a normal reaction and a sign of your body being overprotective.

If we perform a scan (for example x-ray) of your knee, it might not look the same as the knee of someone who does not have pain. This is not always related to how much pain you feel and does not change how you should manage your knee pain.

When you receive the diagnosis Patellar Tendinopathy, the healthcare professional is certain that there is not any serious damage in your knee, and nothing has been missed. Further examinations are not needed to confirm the diagnosis or determine the treatment.

**What is causing my knee pain?**

Children and adolescents who are active in sports are more likely to have pain due to Patellar Tendinopathy.

It can be hard to find the exact reason why you have Patellar Tendinopathy because it is rarely caused by a single event or movement. The balance between activity and rest seems to play an important role.

How much pain you feel depends on several different things that do not always relate directly to your knee. These could be your mood and thoughts or how you use your knee when feeling pain.

Some might worry about their knee pain. Things like running, jumping, and walking on stairs might be painful and some might wonder “should I stop?” or “am I making things worse?”. Worrying about pain, being afraid of hurting your knee, and worrying about how the future will be, can make everything feel worse.

**How do I manage my knee pain?**

Understanding which things affect your pain will help you choose the right treatment for you and help you to control how much pain you feel.

Most adolescents with Patellar Tendinopathy feel that their lives change because their knee hurts. Changes might include how much sport they can take part in. Luckily, you can learn to be physically active even though you have Patellar Tendinopathy. In fact, physical activity is good for you in many ways.

Your body and knee will respond best to small changes in activity. Finding a balance between physical activity and rest may be a good place to start when you have knee pain. It is completely normal to feel more pain sometimes when you are trying to find the right balance.

#### 5. *Patellofemoral Pain (English)*

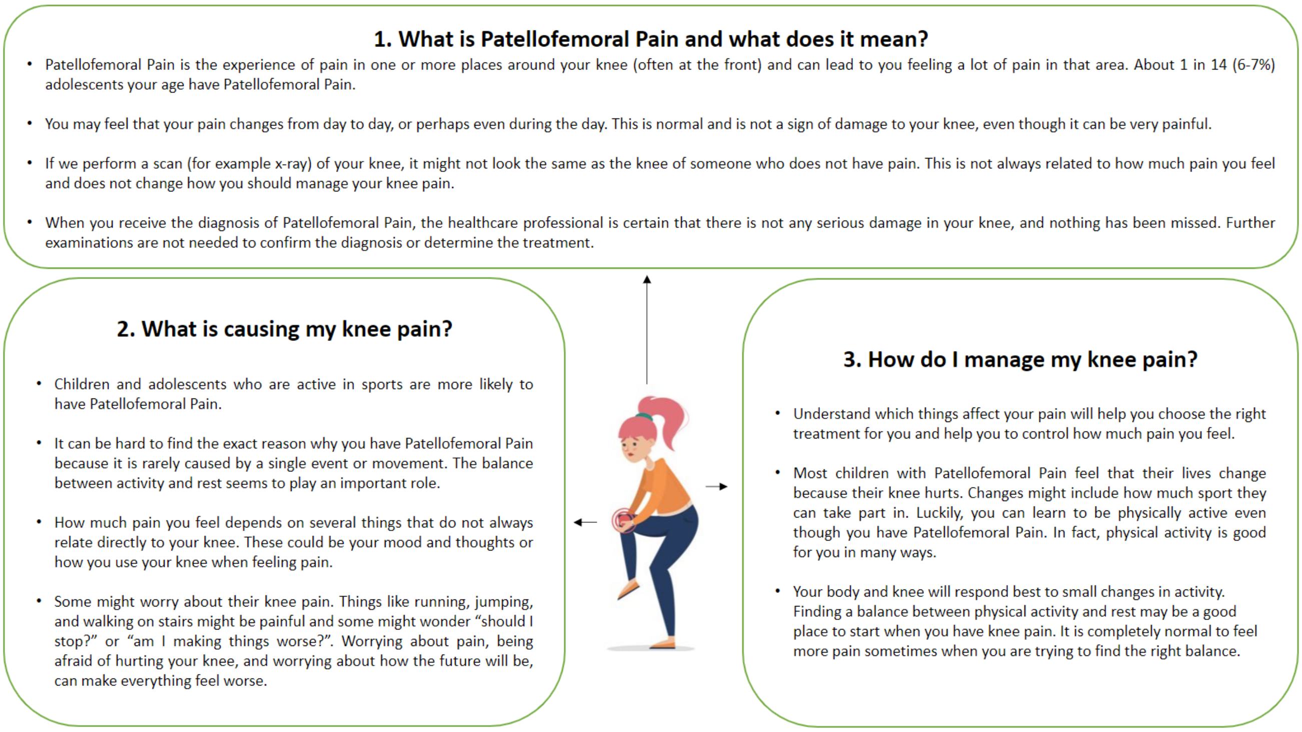

**What is Patellofemoral Pain and what does it mean?**

Patellofemoral Pain is the experience of pain in one or more places around your knee (often at the front), and can lead to you feeling a lot of pain in that area. About 1 in 14 (6-7%) adolescents your age have Patellofemoral Pain.

#### *6. Sinding-Larsson-Johansson (English)*

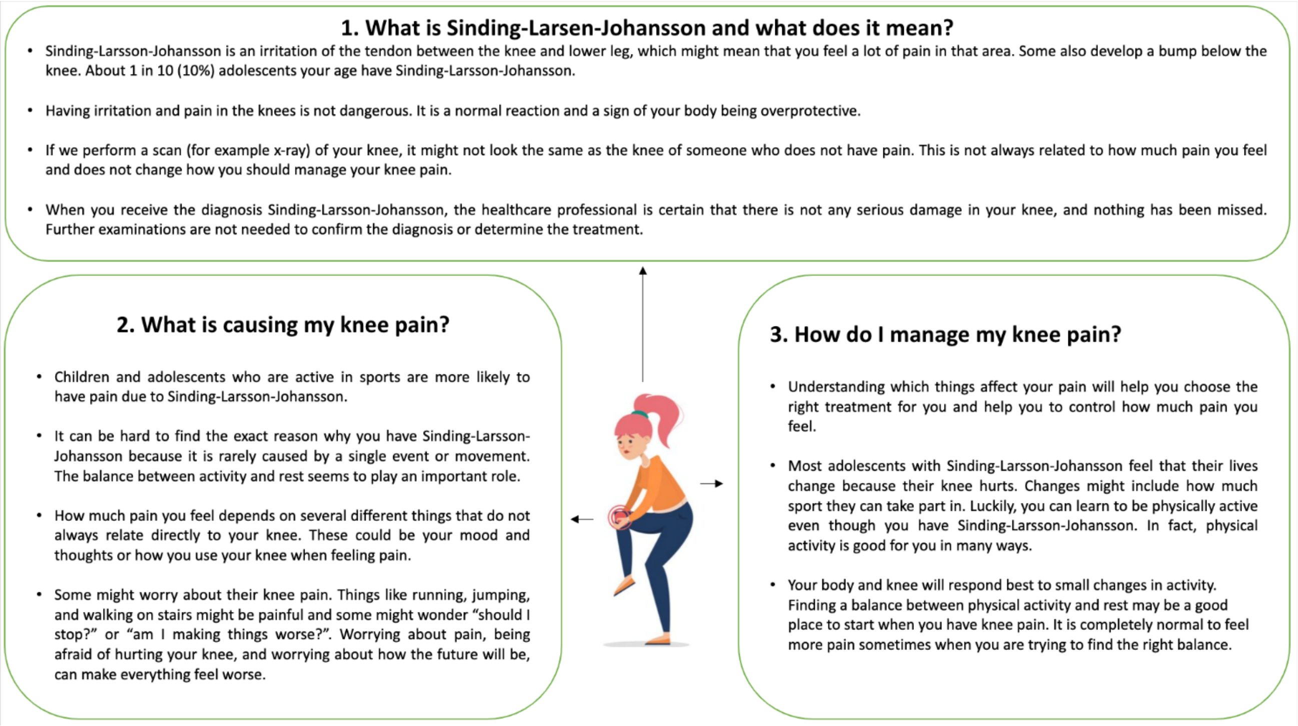

**What is Sinding-Larsson-Johansson and what does it mean?**

Sinding-Larsson-Johansson is an irritation of the tendon between the knee and lower leg, which might mean that you feel a lot of pain in that area. Some also develop a bump below the knee. About 1 in 10 (10%) adolescents your age have Sinding-Larsson-Johansson.

Having irritation and pain in the knees is not dangerous. It is a normal reaction and a sign of your body being overprotective.

If we perform a scan (for example x-ray) of your knee, it might not look the same as the knee of someone who does not have pain. This is not always related to how much pain you feel and does not change how you should manage your knee pain.

When you receive the diagnosis Sinding-Larsson-Johansson, the healthcare professional is certain that there is not any serious damage in your knee, and nothing has been missed. Further examinations are not needed to confirm the diagnosis or determine the treatment.

**What is causing my knee pain?**

Children and adolescents who are active in sports are more likely to have pain due to Sinding-Larsson-Johansson.

It can be hard to find the exact reason why you have Sinding-Larsson-Johansson because it is rarely caused by a single event or movement. The balance between activity and rest seems to play an important role.

How much pain you feel depends on several different things that do not always relate directly to your knee. These could be your mood and thoughts or how you use your knee when feeling pain.

Some might worry about their knee pain. Things like running, jumping, and walking on stairs might be painful and some might wonder “should I stop?” or “am I making things worse?”. Worrying about pain, being afraid of hurting your knee, and worrying about how the future will be, can make everything feel worse.

**How do I manage my knee pain?**

Understanding which things affect your pain will help you choose the right treatment for you and help you to control how much pain you feel.

Most adolescents with Sinding-Larsson-Johansson feel that their lives change because their knee hurts. Changes might include how much sport they can take part in. Luckily, you can learn to be physically active even though you have Sinding-Larsson-Johansson. In fact, physical activity is good for you in many ways.

Your body and knee will respond best to small changes in activity. Finding a balance between physical activity and rest may be a good place to start when you have knee pain. It is completely normal to feel more pain sometimes when you are trying to find the right balance.
